# Revisiting insulin resistance in human cancer cachexia – a systematic review and meta-analysis

**DOI:** 10.1101/2025.03.28.25324822

**Authors:** Jonas Sørensen, Anna Hammershøi, Joan Miquel Màrmol, Louise Lang Lehrskov, Ole Nørgaard, Lykke Sylow

## Abstract

**Background:** Approximately half of all patients with cancer experience unintentional weight loss called cancer cachexia (CAC), which reduces overall survival and impairs quality of life. Metabolic dysfunction is frequently observed in patients with cancer, likely due to insulin resistance. Due to its anabolic effects, insulin resistance could be implicated in the progression of CAC. However, comprehensive clinical data in human populations remain limited, and the potential association between insulin resistance and CAC has yet to be clearly established. Covering this knowledge gap will guide our search for treatable targets to alleviate patients in the future.

**Methods:** To address this knowledge gap, we performed a systematic review and meta-analysis, including comprehensive searches in MEDLINE, Embase, and the Cochrane Central Register of Controlled Trials (CENTRAL). By including studies reporting both fasting levels of insulin and glucose in patients with cancer and CAC according to the internationally accepted CAC definition, we calculated HOMA-IR (Homeostatic Model Assessment for Insulin Resistance) index for each study and thereby estimated the level of insulin resistance (defined as HOMA-IR above 2.0) in patients with CAC. A subgroup analysis was conducted from studies reporting a HOMA-IR index both from a group of patients with CAC and a group of patients without CAC (non-CAC).

**Results:** The mean HOMA-IR of all studies was 1.84 (95%CI: 1.77-1.91). Seventeen studies, with a total of 197 patients with CAC, conducted from 1982 to 2007, fulfilled the inclusion criteria. Age ranged from 52 to 68 years. Five studies reported a HOMA-IR above 2.0, indicative of insulin resistance. Twelve studies found HOMA-IR below 2.0. No differences between disease- or patient characteristics between the studies reporting HOMA-IR values above 2.0 vs. those below 2.0 were identified. Five of the 17 studies also reported HOMA-IR from a group of patients with cancer without CAC, allowing for a separate meta-analysis. In 37 patients with CAC and 49 patients without CAC we documented a mean difference of -0.42 (95%CI: -2.24-1.40) in favor of a lower HOMA-IR in patients with CAC compared to without CAC. Heterogeneity between studies was significant with I^2^ =□94% (P□<□0.05).

**Conclusions:** This systematic review indicates that insulin resistance in patients with cancer and cachexia does not manifest to the same extent as in cancer populations without cachexia. These findings challenge the prevailing understanding of insulin resistance as a driver of cancer cachexia and underscore the need for further longitudinal research in this area.

## Introduction

Cancer-associated weight loss – termed cancer cachexia (CAC) – lowers tolerance to anticancer treatment, reduces quality of life, and impairs survival [1], [2], [3]. CAC is estimated to directly contribute to one in five cancer-related deaths [4], yet no approved drug to treat CAC is available. The underlying metabolic disturbance of CAC is not fully understood but involves the rapid depletion of both adipose tissue and skeletal muscle mass [5], [6], [7]. Insulin is a major inhibitor of adipose tissue lipolysis and controls muscle proteolysis and therefore could play a role in tissue wasting processes. While certain rodent studies indicate that insulin resistance contribute to CAC [8], [9], human data remains scarce, and the potential association between insulin resistance and CAC requires further investigation.

We recently established that patients with various cancers were markedly insulin-resistant [10]. Insulin resistance is a primary defect in type 2 diabetes and accordingly, patients with cancer have an increased risk of new onset type 2 diabetes after their cancer diagnosis [11], [12].

The development of CAC could be related to insulin resistance as preexisting diabetes is related to a greater weight loss in patients with colorectal and pancreatic cancer compared to patients without diabetes [13]. However, that study did not directly assess insulin resistance and thus could not address whether there was an association between insulin resistance and CAC. Another study, that documented insulin resistance in patients with cancer using the gold standard hyper-insulinemic euglycemic clamp method, did not include information on weight loss [10], making it difficult to conclude on the association between insulin resistance and CAC. Other clinical studies have aimed to untangle the role of insulin resistance in CAC using fasting insulin as a surrogate measure, but the findings are inconclusive and inconsistent [14], [15]. It therefore remains an unresolved question whether the documented association between cancer and insulin resistance [10], is related to the development of CAC. Such effort is challenging because the direct measurement of insulin sensitivity in humans requires a hyper-insulinemic euglycemic clamp that is invasive and time consuming. Nonetheless, insulin resistance leads to elevated circulating insulin levels with, or without, an increase in blood glucose levels. These changes can be assessed using the Homeostatic Model Assessment of Insulin Resistance (HOMA-IR). Although HOMA-IR primarily estimates hepatic insulin resistance [16], it serves as a surrogate measure of whole-body insulin resistance [17] and demonstrates correlation with the hyper-insulinemic- euglycemic clamp [18]. Lower HOMA-IR values reflect lower blood glucose levels in relation to circulating insulin, indicating enhanced insulin sensitivity, whereas higher values suggest increased insulin resistance.

To address whether insulin resistance is related to the development of CAC, we undertook a systematic review of studies assessing fasting blood glucose and insulin to calculate HOMA-IR. We found no indications of insulin resistance, challenging the notion that insulin resistance is associated with CAC.

## Method

This systematic review was guided by the Preferred Items for Systematic Review and Meta-Analyses (PRISMA) statement [19] and PRISMA Checklist (Supporting Information S1). A review protocol was published before data extraction and can be accessed on https://www.researchregistry.com (#1869).

### Search strategy

The databases MEDLINE (via Ovid), Embase (via Ovid), and Cochrane Central Register of Controlled Trials (CENTRAL) were searched on 4 June 2024. The search was developed around three concepts: (1) cancer in humans, (2) cachexia and (3) insulin resistance expressed by HOMA-IR index. Using a combination of subject terms from the available controlled vocabularies (Medical Subject Headings and Emtree) as well as free-text terms. No restrictions were applied to publication type or language. The final search string was constructed for MEDLINE and subsequently translated to Embase and CENTRAL (Supporting Information S2a-c) by an information specialist (O.N.). In addition, the reference lists of the included studies were screened (backward citation) as well as studies citing the included studies (forward citation) using the online tool Citation Chaser [20].

### Eligibility criteria

Studies were included based on the following criteria: 1) patients with cancer aged 18 years or older; 2) studies that report data on weight loss and sarcopenia, sufficient for the diagnosis of CAC. The diagnostic criterion for CAC was more than 5% loss of stable body weight over the past 6 months OR a BMI less than 20 kg/m² and ongoing weight loss of more than 2% OR sarcopenia and ongoing weight loss of more than 2% without entering the refractory stage [6]. Sarcopenia is here defined as L3 CT-derived skeletal muscle index (SMI) (cm2/m2) < 43/41 for normal or underweight men/women and <53/41 for overweight and obese men/women [3]; and 3) studies that report data on fasting blood glucose and plasma/serum insulin levels including glucose and plasma/serum insulin levels obtained during an OGTT. Studies were excluded based on the follwing criteria: 1) studies only on non-cancer patients; 2) studies only on patients with hematologic malignancies; 3) studies without specific weight loss data; (4) studies without specific fasting glucose and insulin data; 5) studies with patients with documented diabetes, reported use of anti-diabetic medication or daily use of steroids or no specific information on diabetes or use of antidiabetic drugs; 6) studies including cancer survivors or studies that did not specify the cancer status of the patients; 7) studies reusing data from previously included, thus reanalyzing the same patients; and 8) studies on animals.

### Study selection

To determine whether studies could be assessed further for inclusion, three authors (J.S., A.H., and L.L.L.) independently double-screened the titles and abstracts of all identified records for eligibility. Full-text reports of the remaining studies were double-screened by the same three authors. Discrepancies were resolved through discussions and consensus. The screening process was carried out in EPPI-Reviewer Web [21].

### Data extraction

The following data were extracted from each study and are summarized in Table 1a:

**Table 1.** Characteristics of the included studies. F/M=female/male. SD=standard deviation. NS=not stated. HOMA-IR=Homeostatic Model Assessment of Insulin Resistance

| First author | Year | Country | Cancer site | Patient, n (F/M) | Patient, age mean (SD) | Weight loss (%) | Glucose mean, mmol/L (SD) | Insulin mean, mU/L (SD) | HOMA-IR |
| --- | --- | --- | --- | --- | --- | --- | --- | --- | --- |
| Bennegård K et al. (1) | 1982 | Sweden | Various | 19 (NS) | 59 (±2) | 19% | 5.26 (±0.33) | 7 (±1) | 1.64 |
| Bennegård K et al. (2) | 1983 | Sweden | Various | 8 (NS) | 54 (±6) | 18% | 5.02 (±0.2) | 7 (±1) | 1.56 |
| Burt ME et al. | 1983 | US | Esophageal | 6 (1/5) | 65 (±3) | 19.9% | 3.45 (±0.11) | 5.3 (±1.3) | 0.81 |
| Eden E et al. | 1984 | Sweden | Various | 8 (NS) | 55 (±5) | 15% | 4.92 (±0.2) | 7 (±1) | 1.53 |
| Holroyde c et al. | 1984 | US | Colorectal | 12 (NS) | 68 (NS) | 10% | 5 (±0.12) | 14.9 (±2) | 3.31 |
| Heber D et al. | 1985 | US | Lung | 38 (NS) | 59 (±2) | 16% | 5.16 (±0.1) | 16 (±2) | 3.67 |
| Bennegård K et al. (3) | 1986 | Sweden | Various | 5 (NS) | 60 (±2) | 18% | 4 (±0.61) | 5 (±2) | 0.89 |
| Selberg o et al. | 1990 | UK | Various | 4 (NS) | 56 (NS) | 10% | 4.86 (±1.21) | 4.9 (±1.22) | 1.06 |
| Cersosimo E et al. | 1991 | US | Gastrointestinal | 5 (2/3) | 52 (±5) | 15% | 5.44 (±0.16) | 9 (±2) | 2.18 |
| Heslin M et al. | 1992 | US | Various | 8 (3/5) | 55 (±2) | 18% | 5 (±0.16) | 8 (±1) | 1.78 |
| McCall J et al. | 1992 | New Zealand | Gastrointestinal | 3 (NS) | (NS) | 14.3% | 4.73 (±1.18) | 12.63 (±3.1) | 2.65 |
| Pisters P et al. | 1992 | US | Gastrointestinal | 6 (NS) | 56 (NS) | 17.5% | 5.17 (±0.16) | 11 (±3) | 2.55 |
| Rofo A et al. | 1994 | Australia | Various | 35 (NS) | (NS) | 13% | 5.11 (±0.1) | 7.8 (±0.2) | 1.77 |
| Yoshikawa T et al. | 1994 | Japan | Various | 5 (NS) | (NS) | 8.6% | 4.77 (±0.06) | 8.3 (±3.2) | 1.78 |
| Barber MD et al. | 2000 | UK | Pancreatic | 16 (NS) | 63 (±4) | 17.7% | 5.5 (±0.81) | 3.3 (±2.22) | 0.81 |
| Leij-Halfwerk S et al. | 2000 | The Netherlands | Lung | 9 (NS) | 67 (NS) | 12% | 5.8 (±0.3) | 5.85 (±2) | 1.51 |
| Agustsson T et al. | 2007 | Sweden | Gastrointestinal | 15 (3/12) | 65 (±5) | 20% | 6.4 (±1.6) | 6.7 (±3.3) | 1.90 |

First author (reference), Year of publication, Population country, Target population, Number of eligible patients in the study, Population sex distribution, Population age, Weight loss %, Fasting glucose, Fasting insulin, Calculated HOMA-IR.

### Quality assessment

Two review authors (J.S., A.H.) independently assessed the methodological quality of the included studies using appropriate checklists provided by JBI [22] (Supporting Information S3-19). All eligible studies were included regardless of methodological quality, but their limitations were considered in the interpretation of results [23].

### Data analysis

The research was synthesized in a systematic manner. The data extracted from the reviewed studies was based on: 1) insulin sensitivity (IR) reported as HOMA-IR = (insulin × glucose) / 22.5 for the glucose concentration in mmol/L, or: HOMA-IR = (insulin × glucose) / 405 for glycemia in mg/dL, in both cases, the insulin is in mU/L or uU/ml; and 2) weight loss% (WL%) reported in the most detailed version possible.

Insulin resistance has been defined as a HOMA-IR cut-off of 2.0 representing 25% of the population with the highest fasting insulin concentrations [17], [24]. This cut-off was applied and the prevalence of insulin resistance in the CAC patients was calculated.

HOMA-IR were investigated through standard deviation (SD) with a 95% confidence interval (CI) using the inverse variance method, Cohen statistic, and a random-effects model.

Heterogeneity detected by I square (I2) test, significant heterogeneity was defined as I^2^D>D50% with a PD<D0.05. Subgroup analysis was performed to discover source of heterogeneity. To investigate the influence of each study on pooled effect size we used sensitivity analysis.

## Results

### Eligible studies

Our search yielded 3138 records. After removal of 382 duplicates, 2756 records remained for title and abstract screening (Fig. 1). Following this screening, 52 records remained for full-text screening. It was possible to retrieve all the full-text articles. Thirty-eight records were excluded for not meeting our eligibility criteria (see reasons for exclusion in Supporting Information S20). Additionally, 3 reports were identified through citation searching. In total, 17 studies including 197 patients with cancer and CAC reported measures on fasting glucose and insulin and were included in the analysis. Studies were published from 1982 to 2007.

**Fig. 1.**
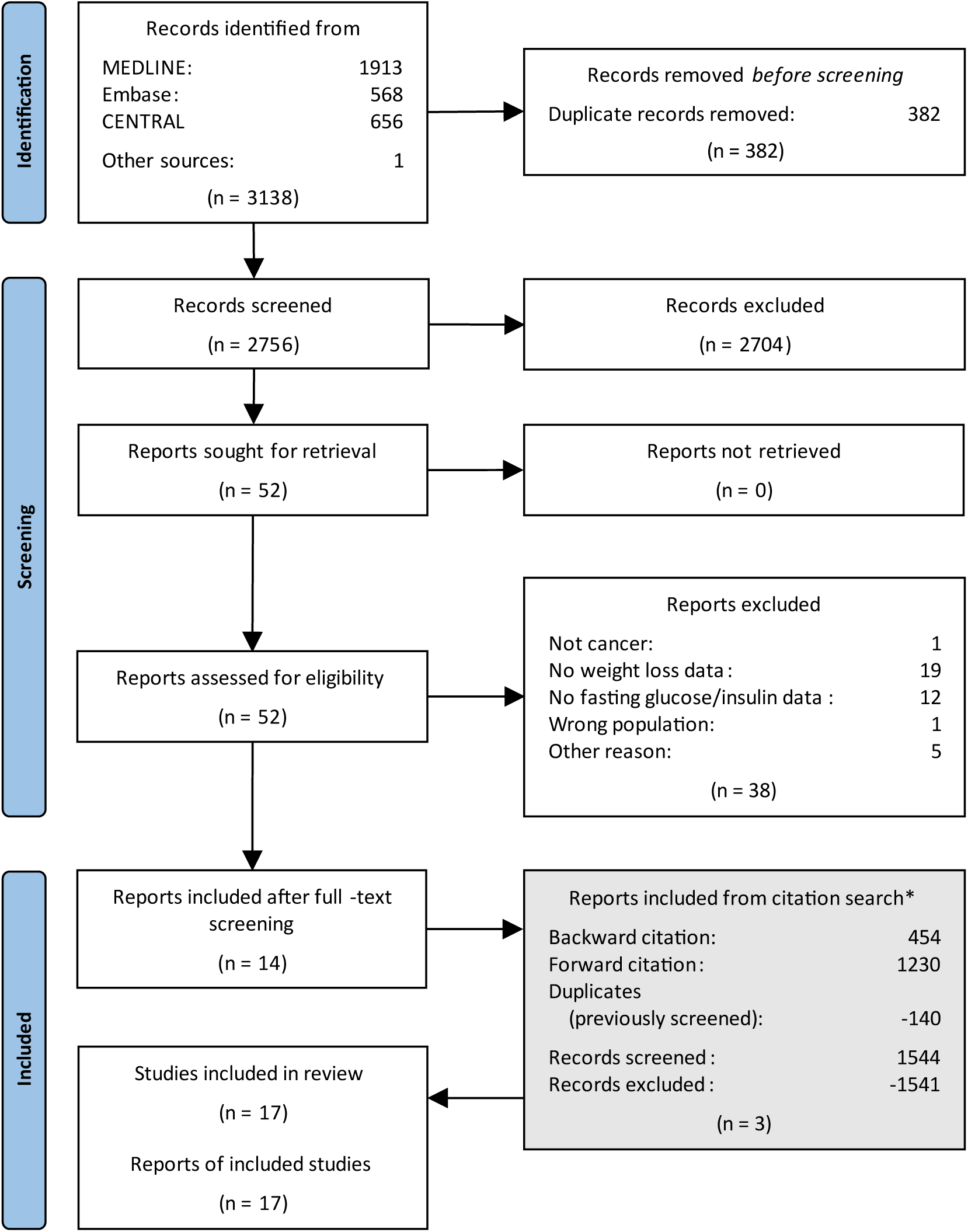
Flow diagram of study selection. *The reports identified from the citation search are located at the bottom to maintain a chronological flow in the diagram. However, they have gone through the same process of deduplication, title and abstract screening, and full-text screening

For all eligible studies, direct personal contact was attempted by contacting the authors via email. In two cases a personal reply from 1^st^ author resulted in individual values from 3 patients [25] and 4 patients [26], respectively. No other authors were able to share individual values on fasting glucose and insulin. For that reason, individual values for each patient in this review were not available and therefore one mean HOMA-IR value was used per study. Studies reporting blood glucose and insulin values from arterial blood levels were accepted [27], [28] due to the fasting and resting condition of the patients, the arterial-venous difference was assumed zero [29]. In 2 studies it was possible from a manuscript figure to determine the mean values of glucose [30] and insulin [31], respectively, by using the tool PlotDigitizer [32]. In these 2 studies the SD were estimated as one-quarter of the mean.

Where glucose and insulin levels were reported with SE (Standard Error) SD was calculated by multiplying SE by the square root of the sample size. One study reported glucose and insulin in median/IQR [33] – here we assumed normal distribution (mean = median) and estimated SD = IQR/1.35. SD for the calculated HOMA-IR could be calculated as the square root of the HOMA-IR variance, and 95%CI = mean ±1.96 x SD/√n.

### Study characteristics

Table 1 summarizes the main characteristics of the included studies. Of the 17 studies included with a total of 197 patients with CAC the HOMA-IR values ranged from 0.81 to 3.67. All studies except 3 [25], [34], [35] reported the mean age, ranging from 52 to 68 years. Five studies reported on the distribution between females and males in patients with CAC [26], [36], [37], [38], [39]. The sample size in each study ranged from 3 to 38 patients. One study [39] included 2 separate groups of patients with CAC – 1 with and 1 without secondary nutrition impact symptoms e.g. anorexia. Without contradicting the inclusion criteria, these 2 CAC groups were merged into 1.

Two studies included solely patients with lung cancer [31], [40]; 7 studies included patients with gastrointestinal cancers, esophageal cancer [36], colorectal cancer [30], upper-gastrointestinal cancer [41], gastro-intestinal cancers not otherwise specified [25], [37], [39], pancreatic cancer [33]; and 8 studies included patients with various cancer diagnoses [14], [15], [27], [28], [34], [35], [38], [42]. Eight studies reported that all included patients were untreated for their cancer [14], [27], [28], [33], [37], [39], [41], [42]. Two studies reported that a fraction of the patients included were pretreated for the cancer [31], [38]. One study reported that included patients did not have any anticancer treatment 3 months prior to the study [25]. One study reported that patients were included immediately prior to the next line treatment [34]. One study stated that all patients were pretreated for the cancer [30]. Eight studies reported that some or all patients had advanced disease [25], [27], [28], [30], [31], [34], [35], [42]. Two studies reported that patients included only had local disease [36], [41]. None of the studies reported clearly about ethnicity in the patients included. All but 1 study [35] were performed in the US, Europe, Australia or New Zealand.

### Meta-analyses

The mean HOMA-IR of the 17 studies with patients with CAC was 1.84 (95%CI: 1.77-1.91). Fig. 2 shows a diagram with individual HOMA-IR and 95%CI depicted along a vertical line representing the cut off value of 2.0 (see Data analysis section). In five studies, the patients had a mean HOMA-IR higher than the defined cut-off of 2.0 [25], [30], [37], [40], [41]. In 12 studies, the patients had a mean HOMA-IR below 2.0; 4 below 1.1 [14], [15], [33], [36]; and in 8 of the studies mean HOMA-IR was between 1.5 and 2.0 [27], [28], [31], [34], [35], [38], [39], [42]. Thus, overall, we did not find indications of insulin resistance in 12 of the 17 included studies, challenging the notion that insulin resistance is associated with CAC.

**Fig. 2.**
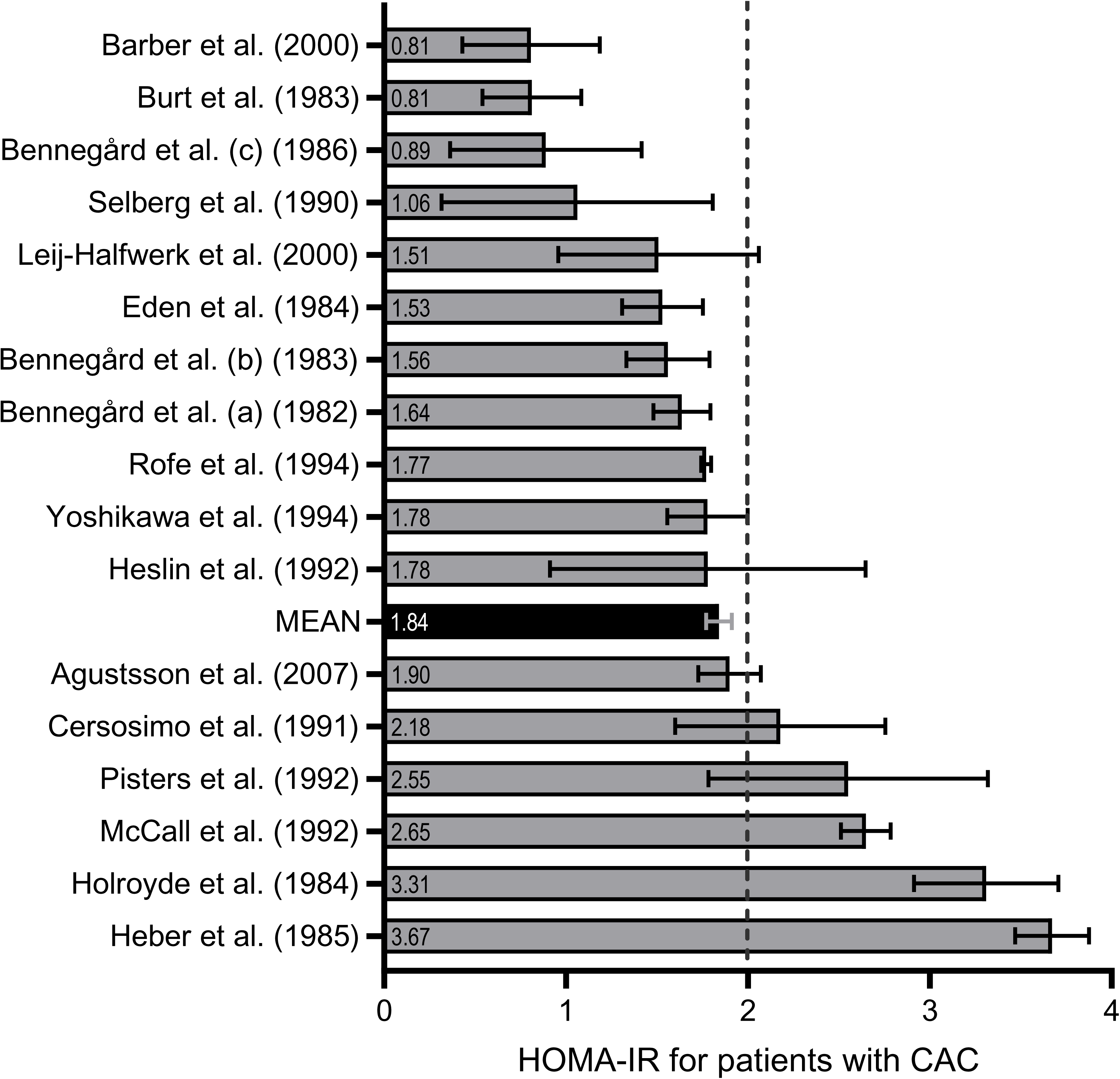
HOMA-IR for studies on patients with cancer cachexia. Studies are lined up according to the individual calculated HOMA-IR values including confidence interval (CI). A vertical dotted line represents the HOMA-IR cut-off value of 2.0. HOMA-IR=Homeostatic Model Assessment of Insulin Resistance. CAC=cancer cachexia

Next, we aimed to compare HOMA-IR between patients with cancer and CAC and those with cancer but without CAC. However, not all studies provided data allowing comparisons with a cancer population without CAC, limiting a direct assessment of the difference in HOMA-IR between the 2 groups. Nevertheless, we identified 5 out of 17 included studies that reported HOMA-IR values for a non-CAC group, defined as having no weight loss or weight loss of <5% in the last 6 months [6] (Table 2). A secondary analysis of these studies was therefore possible [25], [31], [35], [38], [39] and we analyzed HOMA-IR from 5 studies including 37 patients with CAC and 49 patients with non-CAC (Fig. 3). Here we observed a mean difference of -0.42 (95%CI: -2.24-1.40). In the CAC and non-CAC groups, the HOMA-IR index ranged from 1.51 to 2.65 and 0.89 to 4.72, respectively (Fig. 3). Thus, this separate meta-analysis showed a mean difference of -0.42 in favor of a lower HOMA-IR in patients with CAC compared to patients with non-CAC (Fig. 3). This finding aligns with our previous results (Fig. 2) and suggests that CAC is not associated with insulin resistance.

**Fig. 3.**
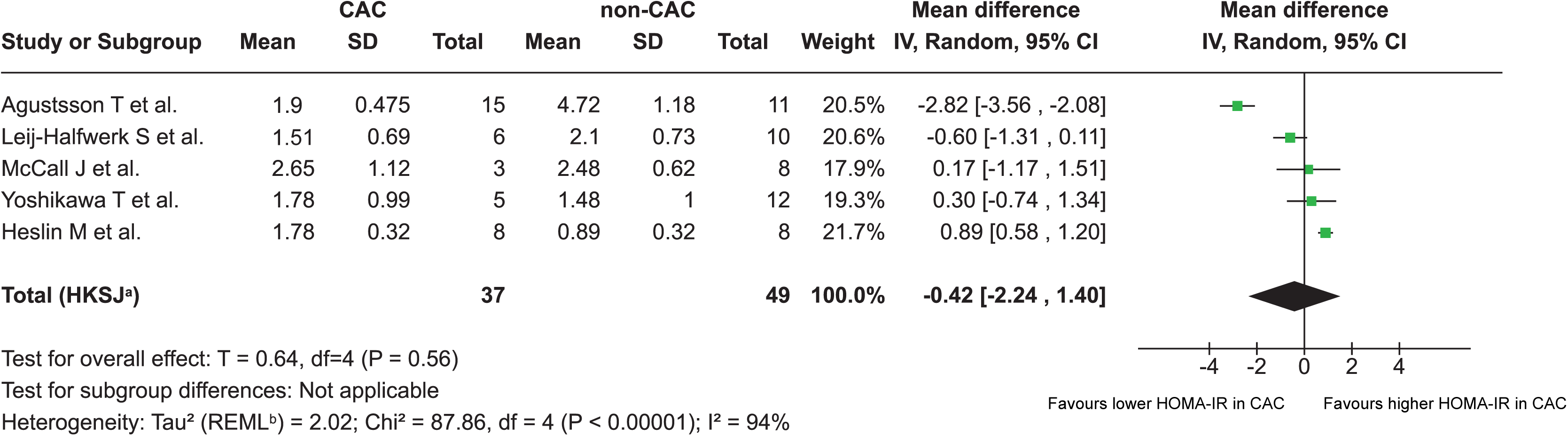
Forest plot comparing HOMA-IR stratifying patients with and without cancer cachexia. Forest plot comparing HOMA-IR in the five studies (mean) stratifying patients with CAC and non-CAC. CAC=patients with cancer cachexia. non-CAC=patients without cancer cachexia. SD=standard deviation. CI=confidence interval. IV=inverse variance. HOMA-IR=Homeostatic Model Assessment of Insulin Resistance. A Random effect model is used to account for both within study and between-study variability. ^a^ CI calculated by Hartung-Knapp-Sidik-Jonkman method. ^b^ Tau^2^ calculated by Restricted Maximum-Likelihood method

**Table 2.** Characteristics of the nonCAC groups, used for subgroup analysis. F/M=female/male. SD=standard deviation. NS=not stated. HOMA-IR: Homeostatic Model Assessment of Insulin Resistance

| First author | Year | Cancer site | Patient, n<br>(F/M) | Glucose<br>mean, mmol/L<br>(SD) | Insulin mean,<br>mU/L (SD) | HOMA-IR |
| --- | --- | --- | --- | --- | --- | --- |
| Heslin M et al. | 1992 | Various | 8 (NS) | 5 (±0.1) | 4 (±1) | **0.89** |
| McCall J et al. | 1992 | Gastrointestinal | 8 (NS) | 4.2 (±0.11) | 13.22 (±8.87) | **2.48** |
| Yoshikawa T et al. | 1994 | Various | 12 (NS) | 5.21 (±0.05) | 6.4 (±3.1) | **1.48** |
| Leij-Halfwerk S et al. | 2000 | Lung | 10 (NS) | 5.3 (±0.2) | 8.92 (±2.23) | **2.10** |
| Agustsson T et al. | 2007 | Gastrointestinal | 11 (NS) | 6.6 (±1.8) | 16.1 (±3.1) | **4.72** |

### Quality assessment

The results of the methodological quality assessment of the studies included using the JBI Critical Appraisal Tools are shown in Supplemental material. We observed significant heterogeneity between the studies with I^2^D=D94% with a PD<D0.05. A funnel plot assessing publication bias was ultimately opted out due to the low sample size of 5 studies in the subgroup analysis.

## Discussion

To our knowledge, this is the first systematic review examining the association between CAC and insulin resistance, assessed by HOMA-IR. A total of 17 studies comprising 197 patients with cancer and CAC were included. Two major findings derived from this study. First, our results indicate that most patients with cancer and CAC are within the normal insulin sensitivity reference range. Second, a sub-analysis of the 5 studies that also included a non-CAC group pointed to a lower HOMA-IR in patients with CAC, indicating less insulin resistance in CAC compared to non-CAC.

Our first finding, that most patients with cancer and CAC fall within the normal insulin sensitivity range, suggests that insulin resistance is not a primary driver of CAC. At first glance, these results appear to contradict previous evidence and the prevailing belief that insulin resistance is associated with CAC. Notably, patients with cancer in general exhibit insulin resistance as documented in a recent meta-analysis [10] and accordingly have a higher probability risk of developing type 2 diabetes [11], [12]. Moreover, patients with cancer have been reported to respond to elevated glucose levels in a diabetic manner with reduced glucose disposal curves [43], [44]. While documenting insulin resistance and dysregulated glucose metabolism, none of those studies stratified patients with CAC, which was done in our study. These present results indicate that CAC is not associated with insulin resistance.

In contrast to the overall normal fasting blood glucose in all but 2 studies (1 reporting hypoglycemia of 3.45 mmol/L [36] and 1 reporting hyperglycemia of 5.8 mmol/L [31]), we observed great variation in fasting insulin levels between studies. There is no international standard normal range to fasting insulinemia in a non-diabetic population, but a normo- insulinemic range from 2-15 mU/L is accepted [24], [45]. Five studies were found in the low-normal end with insulin levels <6 mU/L [14], [15], [31], [33], [36]. Five studies with normal to high insulin levels (≥9 mU/L) were identified, all of which also had a calculated HOMA-IR >2.0.[25], [30], [37], [40], [41].

While most studies reported that patients with cancer and CAC fall within the normal insulin sensitivity range, we did identify 5 studies with a mean HOMA-IR >2.0 [25], [30], [37], [40], [41], indicative of insulin resistance (Fig. 2). In all 5 studies, the high HOMA-IR was driven by normal-high insulinemia, and not hyperglycemia. Hyperinsulinemia in the presence of euglycemia is a hallmark of prediabetes that is primarily due to peripheral insulin resistance of the muscle [16]. Unfortunately, peripheral insulin resistance was not determined in this study but is highly common in the general cancer patient population [10]. No common features in relation to cancer types, treatment status, degree of weight loos, age or sex distribution from the five studies with HOMA-IR >2.0 can be extracted besides all conducted in the US or New Zealand.

Our second finding was that patients with CAC were less insulin resistant compared to patients with non-CAC based on HOMA-IR, which is in line with our first major finding.

These findings align with other studies stratifying between CAC and non-CAC patients. Such studies show that in response to a glucose challenge, the insulin response is lower in patients with cancer and CAC compared to non-CAC [46], [47], indicative of increased insulin sensitivity in patients with CAC. Accordingly, underweight patients with cancer exhibited greater insulin sensitivity compared to their normal-weight counterparts, as estimated in response to intravenous glucose infusion [48].

These findings, along with our present results, suggest that while patients with cancer generally exhibit insulin resistance and metabolic dysregulation, patients with CAC appear to either preserve insulin sensitivity or counteract the insulin-resistance-inducing effects of cancer to some extent.

The biological underlying mechanisms were not identified in the present study but could include several factors.

First, lower insulin resistance in patients with CAC, compared to the general cancer population, may be attributed to malnutrition. Both low body weight and reduced food intake are generally associated with increased insulin sensitivity and low insulin levels in humans [48] and preclinical models [49]. Consistently, five of our included studies reported fasting insulin levels within the low-normal range in patients with CAC [14], [15], [31], [33], [36]. Therefore, the well-documented cancer-associated insulin resistance in humans [10] may be partially counteracted by malnutrition and/or progressive weight loss in patients with CAC.

Second, increased glucose disposal into highly glycolytic tumors may contribute to overall improved whole-body glycemic control, leading to a lower HOMA-IR. However, it remains unclear whether tumors that induce CAC exhibit higher metabolic rates and consequently utilize more glucose compared to non-CAC-inducing tumors.

Third, metabolic reprogramming in tissues undergoing wasting may contribute to the observed absence of insulin resistance in patients with CAC compared to the general cancer population. Cancer and cancer-secreted factors can induce profound metabolic disruptions, as documented in numerous preclinical [9], [50], [51], [52] and clinical studies [53], [54], [55]. In the context of CAC, the activation of catabolic pathways and the disruption of energy homeostasis in adipose and muscle tissue may paradoxically enhance insulin sensitivity or induce glucose uptake via insulin independent pathways [56]. This metabolic reprogramming, characteristic of CAC, could play a key role in mitigating insulin resistance.

This study has several limitations that should be considered when interpreting the findings. The study populations are heterogeneous, with variations in treatment regimens, unreported comorbidities, and concurrent therapies. Additionally, the 17 studies were conducted across different institutions over a span of 25 years, with inconsistencies in insulin assays and sample handling that may have influenced HOMA-IR measurements. Furthermore, individual fasting glucose and insulin levels were unavailable, requiring reliance on mean values to calculate HOMA-IR at the group level for each study. Despite these limitations, this is the first comprehensive analysis of studies measuring HOMA-IR in patients with cancer and CAC, indicating that insulin resistance is less pronounced in CAC compared to non-CAC cancer populations.

Further research, particularly longitudinal human studies directly assessing insulin sensitivity, is essential to uncover the biological mechanisms. As our understanding of CAC-associated metabolic alterations advances, exciting discoveries may reshape our approach to cancer metabolism and its complications.

## Conclusion

Our findings suggest that insulin resistance is less pronounced in patients with cancer and CAC compared to the broader cancer population. These results challenge the current understanding of insulin resistance as a driver of CAC.

Further longitudinal studies are needed to uncover molecular disturbances during treatment, identify potential therapeutic targets, and clarify the complex relationship between insulin resistance and CAC.

## Author contributions

Conceptualization – J.S. and L.S. Screening and data extraction – J.S., A.H., and L.L.L. Formal analysis – J.M.M. and J.S. Methodology – all authors. Supervision – L.S. Validation – all authors. Writing: draft – J.S. and L.S. Writing: review and editing – all authors.

## Data Availability

The data that supports the findings of this study are available from the corresponding author, L.S., upon reasonable request.

## Acknowledgments

L.S. is funded by the Novo Nordisk Foundation, The Danish Cancer Society, The Carlsberg Foundation, and Independent Research Fund Denmark. O.N. is employed at Copenhagen University Hospital – Steno Diabetes Center Copenhagen, a public hospital and research institution under the Capital Region of Denmark, which is partly funded by a grant from the Novo Nordisk Foundation.

## Ethical standards

All studies stated clearly that all patients gave their informed consent prior to their inclusion in the study. This manuscript complies with the Ethical guidelines for authorship and publishing in the Journal of Cachexia, Sarcopenia and Muscle.

## S1 Supporting Information

PRISMA checklist

## S2 Supporting Information

Search strings in MEDLINE, Embase and CENTRAL

## S3-19 Supporting Information

Quality assessment - JBI Critical Appraisal Tools

## S20 Supporting Information

Excluded reports after full-text screening including reason for exclusion

## References

[1] D. S. Mytelka, L. Li, and K. Benoit, “Post-diagnosis weight loss as a prognostic factor in non-small cell lung cancer,” J Cachexia Sarcopenia Muscle, vol. 9, no. 1, pp. 86– 92, Feb. 2018, doi: 10.1002/jcsm.12253.

[2] K. Sánchez-Lara et al., “Association of Nutrition Parameters Including Bioelectrical Impedance and Systemic Inflammatory Response With Quality of Life and Prognosis in Patients With Advanced Non-Small-Cell Lung Cancer: A Prospective Study,” Nutr Cancer, vol. 64, no. 4, pp. 526–534, May 2012, doi: 10.1080/01635581.2012.668744.

[3] L. Martin et al., “Cancer Cachexia in the Age of Obesity: Skeletal Muscle Depletion Is a Powerful Prognostic Factor, Independent of Body Mass Index,” Journal of Clinical Oncology, vol. 31, no. 12, pp. 1539–1547, Apr. 2013, doi: 10.1200/JCO.2012.45.2722.

[4] J. M. Argilés, F. J. López-Soriano, B. Stemmler, and S. Busquets, “Cancer-associated cachexia — understanding the tumour macroenvironment and microenvironment to improve management,” Nat Rev Clin Oncol, vol. 20, no. 4, pp. 250–264, Apr. 2023, doi: 10.1038/s41571-023-00734-5.

[5] B. R. Pryce et al., “Muscle inflammation is regulated by NF-κB from multiple cells to control distinct states of wasting in cancer cachexia,” Cell Rep, vol. 43, no. 11, p. 114925, Nov. 2024, doi: 10.1016/j.celrep.2024.114925.

[6] K. Fearon et al., “Definition and classification of cancer cachexia: an international consensus,” Lancet Oncol, vol. 12, no. 5, pp. 489–495, May 2011, doi: 10.1016/S1470-2045(10)70218-7.

[7] J. Sørensen, “Lung Cancer Cachexia: Can Molecular Understanding Guide Clinical Management?,” Integr Cancer Ther, vol. 17, no. 3, 2018, doi: 10.1177/1534735418781743.

[8] D. R. Miksza et al., “Insulin in combination with pioglitazone prevents advanced cachexia in 256-Walker tumor-bearing rats: effect is greater than treatment alone and is associated with improved insulin sensitivity,” Pharmacological Reports, vol. 75, no. 6, pp. 1571–1587, Dec. 2023, doi: 10.1007/s43440-023-00533-w.

[9] S. H. Raun et al., “Adenosine monophosphate-activated protein kinase is elevated in human cachectic muscle and prevents cancer-induced metabolic dysfunction in mice,” J Cachexia Sarcopenia Muscle, vol. 14, no. 4, pp. 1631–1647, Aug. 2023, doi: 10.1002/jcsm.13238.

[10] J. M. Màrmol et al., “Insulin resistance in patients with cancer: a systematic review and meta-analysis,” Acta Oncol (Madr), pp. 1–8, Apr. 2023, doi: 10.1080/0284186X.2023.2197124.

[11] Y. Hwangbo et al., “Incidence of Diabetes After Cancer Development,” JAMA Oncol, vol. 4, no. 8, p. 1099, Aug. 2018, doi: 10.1001/jamaoncol.2018.1684.

[12] L. Sylow et al., “Incidence of New-Onset Type 2 Diabetes After Cancer: A Danish Cohort Study,” Diabetes Care, vol. 45, no. 6, pp. e105–e106, Jun. 2022, doi: 10.2337/dc22-0232.

[13] A. Chovsepian et al., “Diabetes increases mortality in patients with pancreatic and colorectal cancer by promoting cachexia and its associated inflammatory status,” Mol Metab, vol. 73, p. 101729, Jul. 2023, doi: 10.1016/j.molmet.2023.101729.

[14] K. Bennegard, F. Lundgren, K. Lundholm, K. Bennegård, F. Lundgren, and K. Lundholm, “Mechanisms of insulin resistance in cancer associated malnutrition.,” Clin Physiol, vol. 6, no. 6, pp. 539–547, Jun. 1986, doi: 10.1111/j.1475-097x.1986.tb00787.x.

[15] O. Selberg, D. C. McMillan, T. Preston, H. Carse, A. Shenkin, and H. J. G. Burns, “Palmitate turnover and its response to glucose infusion in weight-losing cancer patients,” Clinical Nutrition, vol. 9, no. 3, pp. 150–156, Jun. 1990, doi: 10.1016/0261-5614(90)90047-V.

[16] M. Matsuda and R. A. DeFronzo, “Insulin sensitivity indices obtained from oral glucose tolerance testing: comparison with the euglycemic insulin clamp.,” Diabetes Care, vol. 22, no. 9, pp. 1462–1470, Sep. 1999, doi: 10.2337/diacare.22.9.1462.

[17] B. Hedblad, P. Nilsson, L. Janzon, and G. Berglund, “Relation between insulin resistance and carotid intima-media thickness and stenosis in non-diabetic subjects. Results from a cross-sectional study in Malmö, Sweden,” Diabetic Medicine, vol. 17, no. 4, pp. 299–307, Apr. 2000, doi: 10.1046/j.1464-5491.2000.00280.x.

[18] E. Bonora et al., “Homeostasis model assessment closely mirrors the glucose clamp technique in the assessment of insulin sensitivity: studies in subjects with various degrees of glucose tolerance and insulin sensitivity.,” Diabetes Care, vol. 23, no. 1, pp. 57–63, Jan. 2000, doi: 10.2337/diacare.23.1.57.

[19] M. J. Page et al., “The PRISMA 2020 statement: an updated guideline for reporting systematic reviews,” BMJ, p. n71, Mar. 2021, doi: 10.1136/bmj.n71.

[20] N. R. Haddaway, M. J. Grainger, and C. T. Gray, “Citationchaser: A tool for transparent and efficient forward and backward citation chasing in systematic searching,” Res Synth Methods, vol. 13, no. 4, pp. 533–545, Jul. 2022, doi: 10.1002/jrsm.1563.

[21] J. , G. S., B. J. , G. Z. , O. P. , & B. M. & K. A. Thomas, “EPPI-Reviewer: advanced software for systematic reviews, maps and evidence synthesis.,” *EPPI Centre, UCL Social Research Institute*, University College London, 2023.

22. “https://jbi.global/critical-appraisal-tools.”

[23] Higgins JPT, Thomas J, Chandler J, Cumpston M, Li T, Page MJ, Welch VA (editors). Cochrane Handbook for Systematic Reviews of Interventions version 6.5 (updated August 2024). Cochrane, 2024. Available from www.training.cochrane.org/handbook.

[24] B Balkau and M A Charles, “Comment on the provisional report from the WHO consultation,” Diabetic Medicine, vol. 16, no. 5, pp. 442–443, May 1999, doi: 10.1046/j.1464-5491.1999.00059.x.

[25] J. L. McCall, J. A. Tuckey, and B. R. Parry, “Serum tumour necrosis factor alpha and insulin resistance in gastrointestinal cancer.,” British Journal of Surgery, vol. 79, no. 12, pp. 1361–1363, Dec. 1992, doi: 10.1002/bjs.1800791240.

[26] O. Selberg, D. C. McMillan, T. Preston, H. Carset, A. Shenkin, and H. J. G Burns, “Palmitate Turnover and its Response to Glucose Infusion in Weight-losing Can&r Patients,” Clinical Nutrition, vol. 9, pp. 150–56, 1990.

[27] K. Bennegard, E. Edã©n, L. Ekman, T. Scherstã©n, and K. Lundholm2, “Metabolic Balance across the Leg in Weight-losing Cancer Patients Compared to Depleted Patients without Cancer1,” 1982. [Online]. Available: http://aacrjournals.org/cancerres/article-pdf/42/10/4293/2411671/cr0420104293.pdf

[28] K. Bennegård, E. Edén, L. Ekman, T. Scherstén, and K. Lundholm, “Metabolic Response of Whole Body and Peripheral Tissues to Enteral Nutrition in Weight-Losing Cancer and Noncancer Patients,” Gastroenterology, vol. 85, no. 1, pp. 92–99, 1983, doi: 10.1016/S0016-5085(83)80234-0.

[29] D. M. Bell, “Femoral arteriovenous sugar differences in fasting human beings.,” J Lab Clin Med, vol. 40, no. 3, pp. 337–41, Sep. 1952.

[30] C. P. Holroyde, C. L. Skutches, G. Boden, G. A. Reichard, and and GeorgeA Reichard, “Glucose metabolism in cachectic patients with colorectal cancer.,” Cancer Res, vol. 44, no. 12 Pt 1, pp. 5910–5913, Apr. 1984, [Online]. Available: http://ovidsp.ovid.com/ovidweb.cgi?T=JS&PAGE=reference&D=med2&NEWS=N&AN=6388829

[31] S. Leij-Halfwerk et al., “Altered hepatic gluconeogenesis during L-alanine infusion in weight-losing lung cancer patients as observed by phosphorus magnetic resonance spectroscopy and turnover measurements.,” Cancer Res, vol. 60, no. 3, pp. 618–623, Apr. 2000, [Online]. Available: http://ovidsp.ovid.com/ovidweb.cgi?T=JS&PAGE=reference&D=med4&NEWS=N&AN=10676645

[32] “https://plotdigitizer.com/app.”

[33] M. D. Barber, D. C. McMillan, T. Preston, J. A. Ross, and K. C. Fearon, “Metabolic response to feeding in weight-losing pancreatic cancer patients and its modulation by a fish-oil-enriched nutritional supplement.,” Comment in: Clin Sci (Lond). 2000 Apr;98(4):365 PMID: 10731468 [https://www.ncbi.nlm.nih.gov/pubmed/10731468], vol. 98, no. 4, pp. 389–399, Apr. 2000, [Online]. Available: http://ovidsp.ovid.com/ovidweb.cgi?T=JS&PAGE=reference&D=med4&NEWS=N&AN=10731472

[34] A. M. Rofe, C. S. Bourgeois, P. Coyle, A. Taylor, and E. A. Abdi, “Altered insulin response to glucose in weight-losing cancer patients.,” Anticancer Res, vol. 14, no. 2B, pp. 647–650, Apr. 1994, [Online]. Available: http://ovidsp.ovid.com/ovidweb.cgi?T=JS&PAGE=reference&D=med3&NEWS=N&AN=8010722

[35] T. Yoshikawa, Y. Noguchi, A. Matsumoto, and C. Tekst, “Effects of tumor removal and body weight loss on insulin resistance in patients with cancer.,” Surgery, vol. 116, no. 1, pp. 62–66, Apr. 1994, [Online]. Available: http://ovidsp.ovid.com/ovidweb.cgi?T=JS&PAGE=reference&D=med3&NEWS=N&AN=8023270

[36] M. E. Burt, T. T. Aoki, C. M. Gorschboth, and M. F. Brennan, “Peripheral tissue metabolism in cancer-bearing man.,” Ann Surg, vol. 198, no. 6, pp. 685–691, Apr. 1983, doi: 10.1097/00000658-198312000-00003.

[37] E. Cersosimo et al., “The effect of graded doses of insulin on peripheral glucose uptake and lactate release in cancer cachexia.,” Surgery, vol. 109, no. 4, pp. 459– 467, Apr. 1991, [Online]. Available: http://ovidsp.ovid.com/ovidweb.cgi?T=JS&PAGE=reference&D=med3&NEWS=N&AN=2008651

[38] M. J. Heslin, E. Newman, R. F. Wolf, P. W. Pisters, and M. F. Brennan, “Effect of systemic hyperinsulinemia in cancer patients.,” Cancer Res, vol. 52, no. 14, pp. 3845– 3850, Apr. 1992, [Online]. Available: http://ovidsp.ovid.com/ovidweb.cgi?T=JS&PAGE=reference&D=med3&NEWS=N&AN=1617658

[39] T. Agustsson et al., “Mechanism of Increased Lipolysis in Cancer Cachexia,” Cancer Res, vol. 67, no. 11, pp. 5531–5537, Jun. 2007, doi: 10.1158/0008-5472.CAN-06-4585.

[40] D. Heber, L. O. Byerly, and R. T. Chlebowski, “Metabolic abnormalities in the cancer patient.,” Cancer, vol. 55, no. 1 Suppl, pp. 225–229, Apr. 1985, doi: 10.1002/1097-0142(19850101)55:1+<225::aid-cncr2820551304>3.0.co;2-7.

[41] P. W. Pisters, E. Cersosimo, A. Rogatko, and M. F. Brennan, “Insulin action on glucose and branched-chain amino acid metabolism in cancer cachexia: differential effects of insulin.,” Surgery, vol. 111, no. 3, pp. 301–310, Apr. 1992, [Online]. Available: http://ovidsp.ovid.com/ovidweb.cgi?T=JS&PAGE=reference&D=med3&NEWS=N&AN=1542855

[42] E. Eden et al., “Glucose flux in relation to energy expenditure in malnourished patients with and without cancer during periods of fasting and feeding.,” Cancer Res, vol. 44, no. 4, pp. 1718–1724, Apr. 1984, [Online]. Available:http://ovidsp.ovid.com/ovidweb.cgi?T=JS&PAGE=reference&D=med2&NEWS=N&AN=6367972

[43] D. Heber, R. T. Chlebowski, D. E. Ishibashi, J. N. Herrold, and J. B. Block, “Abnormalities in glucose and protein metabolism in noncachectic lung cancer patients.,” Cancer Res, vol. 42, no. 11, pp. 4815–9, Nov. 1982.

[44] J. A. Norton, M. Maher, R. Wesley, D. White, and M. F. Brennan, “Glucose intolerance in sarcoma patients,” Cancer, vol. 54, no. 12, pp. 3022–3027, Dec. 1984, doi: 10.1002/1097-0142(19841215)54:12<3022::AID-CNCR2820541234>3.0.CO;2-K.

[45] C. Li, E. S. Ford, L. C. McGuire, A. H. Mokdad, R. R. Little, and G. M. Reaven, “Trends in Hyperinsulinemia Among Nondiabetic Adults in the U.S.,” Diabetes Care, vol. 29, no. 11, pp. 2396–2402, Nov. 2006, doi: 10.2337/dc06-0289.

[46] B. Jasani, L. J. Donaldson, J. G. Ratcliffe, and G. S. Sokhi, “Mechanism of impaired glucose tolerance in patients with neoplasia,” Br J Cancer, vol. 38, no. 2, pp. 287– 292, Aug. 1978, doi: 10.1038/bjc.1978.200.

[47] K. Lundholm, G. Holm, and T. Scherstén, “Insulin resistance in patients with cancer.,” Cancer Res, vol. 38, no. 12, pp. 4665–70, Dec. 1978.

[48] J. A. Tayek, S. Manglik, and E. Abemayor, “Insulin secretion, glucose production, and insulin sensitivity in underweight and normal-weight volunteers, and in underweight and normal-weight cancer patients: A Clinical Research Center Study,” Metabolism, vol. 46, no. 2, pp. 140–145, Feb. 1997, doi: 10.1016/S0026-0495(97)90291-2.

[49] C. J. Grace, I. Swenne, P. G. Kohn, A. J. Strain, and R. D. Milner, “Protein-energy malnutrition induces changes in insulin sensitivity.,” Diabete Metab, vol. 16, no. 6, pp. 484–91, Dec. 1990.

[50] S. K. Das et al., “Adipose Triglyceride Lipase Contributes to Cancer-Associated Cachexia,” Science *(1979)*, vol. 333, no. 6039, pp. 233–238, Jul. 2011, doi: 10.1126/science.1198973.

[51] P. Morigny et al., “High levels of modified ceramides are a defining feature of murine and human cancer cachexia,” J Cachexia Sarcopenia Muscle, vol. 11, no. 6, pp. 1459–1475, Dec. 2020, doi: 10.1002/jcsm.12626.

[52] X. Han et al., “Cancer causes metabolic perturbations associated with reduced insulin-stimulated glucose uptake in peripheral tissues and impaired muscle microvascular perfusion,” Metabolism, vol. 105, p. 154169, Apr. 2020, doi: 10.1016/j.metabol.2020.154169.

[53] C. M. Op den Kamp et al., “Nuclear transcription factor κ B activation and protein turnover adaptations in skeletal muscle of patients with progressive stages of lung cancer cachexia,” Am J Clin Nutr, vol. 98, no. 3, pp. 738–748, Sep. 2013, doi: 10.3945/ajcn.113.058388.

[54] M. S. Yule et al., “Biomarker endpoints in cancer cachexia clinical trials: Systematic Review 5 of the cachexia endpoint series,” J Cachexia Sarcopenia Muscle, May 2024, doi: 10.1002/jcsm.13491.

[55] H. A. Ebhardt et al., “Comprehensive proteome analysis of human skeletal muscle in cachexia and sarcopenia: a pilot study,” J Cachexia Sarcopenia Muscle, vol. 8, no. 4, pp. 567–582, Aug. 2017, doi: 10.1002/jcsm.12188.

[56] H. T. Langer, M. Rohm, M. D. Goncalves, and L. Sylow, “AMPK as a mediator of tissue preservation: time for a shift in dogma?,” Nat Rev Endocrinol, vol. 20, no. 9, pp. 526–540, Sep. 2024, doi: 10.1038/s41574-024-00992-y.

