## Supplementary material for "Revisiting insulin resistance in human cancer cachexia – a systematic review and meta-analysis": S1

| **Section and Topic** | **Item #** | **Checklist item** | **Location where item is reported** |
| --- | --- | --- | --- |
| **TITLE** | | |  |
| Title | 1 | Identify the report as a systematic review. | Title page |
| **ABSTRACT** | | |  |
| Abstract | 2 | See the PRISMA 2020 for Abstracts checklist. | Abstract section |
| **INTRODUCTION** | | |  |
| Rationale | 3 | Describe the rationale for the review in the context of existing knowledge. | Introduction section |
| Objectives | 4 | Provide an explicit statement of the objective(s) or question(s) the review addresses. | Introduction section |
| **METHODS** | | |  |
| Eligibility criteria | 5 | Specify the inclusion and exclusion criteria for the review and how studies were grouped for the syntheses. | Methods section |
| Information sources | 6 | Specify all databases, registers, websites, organisations, reference lists and other sources searched or consulted to identify studies. Specify the date when each source was last searched or consulted. | Methods section |
| Search strategy | 7 | Present the full search strategies for all databases, registers and websites, including any filters and limits used. | Methods section |
| Selection process | 8 | Specify the methods used to decide whether a study met the inclusion criteria of the review, including how many reviewers screened each record and each report retrieved, whether they worked independently, and if applicable, details of automation tools used in the process. | ‘Study selection’ in Methods section |
| Data collection process | 9 | Specify the methods used to collect data from reports, including how many reviewers collected data from each report, whether they worked independently, any processes for obtaining or confirming data from study investigators, and if applicable, details of automation tools used in the process. | ‘Data extraction’ in Methods section |
| Data items | 10a | List and define all outcomes for which data were sought. Specify whether all results that were compatible with each outcome domain in each study were sought (e.g. for all measures, time points, analyses), and if not, the methods used to decide which results to collect. | ‘Data analysis’ in Methods section |
|  | 10b | List and define all other variables for which data were sought (e.g. participant and intervention characteristics, funding sources). Describe any assumptions made about any missing or unclear information. | Methods section |
| Study risk of bias assessment | 11 | Specify the methods used to assess risk of bias in the included studies, including details of the tool(s) used, how many reviewers assessed each study and whether they worked independently, and if applicable, details of automation tools used in the process. | ‘Quality assessment’ in Methods section |
| Effect measures | 12 | Specify for each outcome the effect measure(s) (e.g. risk ratio, mean difference) used in the synthesis or presentation of results. | ‘Data analysis’ in Methods section |
| Synthesis methods | 13a | Describe the processes used to decide which studies were eligible for each synthesis (e.g. tabulating the study intervention characteristics and comparing against the planned groups for each synthesis (item #5)). | ‘Study selection’ in Methods section |
|  | 13b | Describe any methods required to prepare the data for presentation or synthesis, such as handling of missing summary statistics, or data conversions. | ‘Data analysis’ in Methods section + ‘Eligible studies’ in Results section |
|  | 13c | Describe any methods used to tabulate or visually display results of individual studies and syntheses. | ‘Data analysis’ in Methods section |
|  | 13d | Describe any methods used to synthesize results and provide a rationale for the choice(s). If meta-analysis was performed, describe the model(s), method(s) to identify the presence and extent of statistical heterogeneity, and software package(s) used. | ‘Data analysis’ in Methods section |
|  | 13e | Describe any methods used to explore possible causes of heterogeneity among study results (e.g. subgroup analysis, meta-regression). | ‘Data analysis’ in Methods section |
|  | 13f | Describe any sensitivity analyses conducted to assess robustness of the synthesized results. | ‘Data analysis’ in Methods section |
| Reporting bias assessment | 14 | Describe any methods used to assess risk of bias due to missing results in a synthesis (arising from reporting biases). | ‘Quality assessment’ in Methods section |
| Certainty assessment | 15 | Describe any methods used to assess certainty (or confidence) in the body of evidence for an outcome. | ‘Data analysis’ in Methods section |
| **RESULTS** | | |  |
| Study selection | 16a | Describe the results of the search and selection process, from the number of records identified in the search to the number of studies included in the review, ideally using a flow diagram. | ‘Eligible studies’ in Results section |
|  | 16b | Cite studies that might appear to meet the inclusion criteria, but which were excluded, and explain why they were excluded. | Supporting Information S20 |
| Study characteristics | 17 | Cite each included study and present its characteristics. | Table 1A, ‘Study characteristics' in Results section |
| Risk of bias in studies | 18 | Present assessments of risk of bias for each included study. | Supporting information S3-19 |
| Results of individual studies | 19 | For all outcomes, present, for each study: (a) summary statistics for each group (where appropriate) and (b) an effect estimate and its precision (e.g. confidence/credible interval), ideally using structured tables or plots. | Figure 2 and Figure 3 in Results section |
| Results of syntheses | 20a | For each synthesis, briefly summarise the characteristics and risk of bias among contributing studies. | ‘Study characteristics’ in Results section |
|  | 20b | Present results of all statistical syntheses conducted. If meta-analysis was done, present for each the summary estimate and its precision (e.g. confidence/credible interval) and measures of statistical heterogeneity. If comparing groups, describe the direction of the effect. | ‘Meta analyses and Figure 3 in Results section |
|  | 20c | Present results of all investigations of possible causes of heterogeneity among study results. | ‘Meta analyses’ and Figure 3 in Results section |
|  | 20d | Present results of all sensitivity analyses conducted to assess the robustness of the synthesized results. | Figure 3 in Results section |
| Reporting biases | 21 | Present assessments of risk of bias due to missing results (arising from reporting biases) for each synthesis assessed. | Supporting information S3-19 |
| Certainty of evidence | 22 | Present assessments of certainty (or confidence) in the body of evidence for each outcome assessed. | Supporting information S3-19 |
| **DISCUSSION** | | |  |
| Discussion | 23a | Provide a general interpretation of the results in the context of other evidence. | Discussion section |
|  | 23b | Discuss any limitations of the evidence included in the review. | Discussion section |
|  | 23c | Discuss any limitations of the review processes used. | Discussion section |
|  | 23d | Discuss implications of the results for practice, policy, and future research. | Conclusion section |
| **OTHER INFORMATION** | | |  |
| Registration and protocol | 24a | Provide registration information for the review, including register name and registration number, or state that the review was not registered. | Methods section |
|  | 24b | Indicate where the review protocol can be accessed, or state that a protocol was not prepared. | Methods section |
|  | 24c | Describe and explain any amendments to information provided at registration or in the protocol. | Methods section |
| Support | 25 | Describe sources of financial or non-financial support for the review, and the role of the funders or sponsors in the review. | Acknowledgments section |
| Competing interests | 26 | Declare any competing interests of review authors. | Disclosure statement |
| Availability of data, code and other materials | 27 | Report which of the following are publicly available and where they can be found: template data collection forms; data extracted from included studies; data used for all analyses; analytic code; any other materials used in the review. | Data extracted from included studies; data used for all analyses; and data that supports the findings of this study. See also Data availability statement |

*From:* Page MJ, McKenzie JE, Bossuyt PM, Boutron I, Hoffmann TC, Mulrow CD, et al. The PRISMA 2020 statement: an updated guideline for reporting systematic reviews. BMJ 2021;372:n71. doi: 10.1136/bmj.n7

Revisiting insulin resistance in human cancer cachexia – a systematic review and meta-analysis

Journal of Cachexia, Sarcopenia and Muscle

Authors

Jonas Sørensena, Anna Hammershøia, Joan Miguel Màrmolb, Louise Lang Lehrskovc, d, Ole Nørgaarde, and Lykke Sylowa*.

Affiliations

a.	Department of Biomedical Sciences, Faculty of Health and Medical Sciences, University of Copenhagen, Copenhagen, Denmark

b.	Department of Nutrition, Exercise, and Sports, Faculty of Science, University of Copenhagen, Copenhagen, Denmark

c.	Center for Physical Activity Research (CFAS), Centre for Cancer and Organ Diseases, Copenhagen University Hospital – Rigshospitalet, Copenhagen, Denmark

d.	Department of Oncology, Copenhagen University Hospital – Herlev and Gentofte, Herlev, Denmark

e.	Department of Education, Danish Diabetes Knowledge Center, Copenhagen University Hospital – Steno Diabetes Center Copenhagen, Herlev, Denmark
