## Supplementary material for "Revisiting insulin resistance in human cancer cachexia – a systematic review and meta-analysis": S2

### Database searches

**Table S2a: Search in MEDLINE (Ovid)**

Date of search: 4 June 2024

| **#** | **Searches** | **Results** |
| --- | --- | --- |
| 1 | exp Neoplasms/ | 3979014 |
| 2 | (adenocarcinoma$ or cancer$ or carcinoma$ or choricarcinoma$ or malignan$ or metasta$ or neoplasm$ or sarcoma$ or teratoma$ or tumo?r$).ti,ab,kf,kw. | 4281894 |
| 3 | or/1-2 | 5317591 |
| 4 | exp Malnutrition/ | 138265 |
| 5 | exp Muscular Atrophy/ | 23541 |
| 6 | exp Weight Loss/ | 51417 |
| 7 | (cachex$ or malnourish$ or malnutri$ or (musc$ adj (atroph$ or loss)) or sarcopeni$ or underweight or wasting or (weight adj3 (loss or redu$))).ti,ab,kf,kw. | 283421 |
| 8 | or/4-7 | 413149 |
| 9 | Blood Glucose/ and (fasting or toleran$).ti,ab,kf,kw. | 52380 |
| 10 | Glucose Clamp Technique/ | 5757 |
| 11 | exp Insulin Resistance/ | 102705 |
| 12 | (((bloodglucose or glucose) adj3 (fasting or toleran$)) or (euglyc?emic adj3 clamp$) or homa-ir or (insulin adj3 (resistan$ or sensitiv$)) or ogtt or ((plasma$ or serum) adj insulin)).ti,ab,kf,kw. | 216902 |
| 13 | or/9-12 | 257298 |
| 14 | and/3,8,13 | 2200 |
| 15 | exp Animals/ not Humans/ | 5227826 |
| 16 | 14 not 15 | 1913 |

Revisiting insulin resistance in human cancer cachexia – a systematic review and meta-analysis

Journal of Cachexia, Sarcopenia and Muscle

Authors

Jonas Sørensena, Anna Hammershøia, Joan Miguel Màrmolb, Louise Lang Lehrskovc, d, Ole Nørgaarde, and Lykke Sylowa*.

Affiliations

a.	Department of Biomedical Sciences, Faculty of Health and Medical Sciences, University of Copenhagen, Copenhagen, Denmark

b.	Department of Nutrition, Exercise, and Sports, Faculty of Science, University of Copenhagen, Copenhagen, Denmark

c.	Center for Physical Activity Research (CFAS), Centre for Cancer and Organ Diseases, Copenhagen University Hospital – Rigshospitalet, Copenhagen, Denmark

d.	Department of Oncology, Copenhagen University Hospital – Herlev and Gentofte, Herlev, Denmark

e.	Department of Education, Danish Diabetes Knowledge Center, Copenhagen University Hospital – Steno Diabetes Center Copenhagen, Herlev, Denmark

**Table S2b: Search in Embase (Ovid)**

Date of search: 4 June 2024

| **#** | **Searches** | **Results** |
| --- | --- | --- |
| 1 | exp neoplasms/ | 5768417 |
| 2 | (adenocarcinoma$ or cancer$ or carcinoma$ or choricarcinoma$ or malignan$ or metasta$ or neoplasm$ or sarcoma$ or teratoma$ or tumo?r$).ti,ab,kf,kw. | 5697222 |
| 3 | or/1-2 | 7053235 |
| 4 | exp malnutrition/ | 200458 |
| 5 | exp muscular atrophy/ | 63682 |
| 6 | exp body weight loss/ | 102050 |
| 7 | (cachex$ or malnourish$ or malnutri$ or (musc$ adj (atroph$ or loss)) or sarcopeni$ or underweight or wasting or (weight adj3 (loss or redu$))).ti,ab,kf,kw. | 424587 |
| 8 | or/4-7 | 603132 |
| 9 | exp blood glucose/ and (fasting or toleran$).ti,ab,kf,kw. | 106230 |
| 10 | exp glucose clamp technique/ | 5137 |
| 11 | insulin resistance/ | 154091 |
| 12 | (((bloodglucose or glucose) adj3 (fasting or toleran$)) or (euglyc?emic adj3 clamp$) or homa-ir or (insulin adj3 (resistan$ or sensitiv$)) or ogtt or ((plasma$ or serum) adj insulin)).ti,ab,kf,kw. | 317319 |
| 13 | or/9-12 | 366596 |
| 14 | and/3,8,13 | 3659 |
| 15 | (rat or rats or mouse or mice or swine or porcine or murine or sheep or lambs or pigs or piglets or rabbit or rabbits or cat or cats or dog or dogs or cattle or bovine or monkey or monkeys or trout or marmoset$1).ti. and animal experiment/ | 1254241 |
| 16 | animal experiment/ not (human experiment/ or human/) | 2637299 |
| 17 | 15 or 16 | 2711386 |
| 18 | 14 not 17 | 3224 |
| 19 | limit 18 to exclude medline journals | 568 |

Revisiting insulin resistance in human cancer cachexia – a systematic review and meta-analysis

Journal of Cachexia, Sarcopenia and Muscle

Authors

Jonas Sørensena, Anna Hammershøia, Joan Miguel Màrmolb, Louise Lang Lehrskovc, d, Ole Nørgaarde, and Lykke Sylowa*.

Affiliations

a.	Department of Biomedical Sciences, Faculty of Health and Medical Sciences, University of Copenhagen, Copenhagen, Denmark

b.	Department of Nutrition, Exercise, and Sports, Faculty of Science, University of Copenhagen, Copenhagen, Denmark

c.	Center for Physical Activity Research (CFAS), Centre for Cancer and Organ Diseases, Copenhagen University Hospital – Rigshospitalet, Copenhagen, Denmark

d.	Department of Oncology, Copenhagen University Hospital – Herlev and Gentofte, Herlev, Denmark

e.	Department of Education, Danish Diabetes Knowledge Center, Copenhagen University Hospital – Steno Diabetes Center Copenhagen, Herlev, Denmark

**Table S2c: Search in CENTRAL (Cochrane Library)**

Date of search: 4 June 2024

| **#** | **Searches (in Trials)** | **Results** |
| --- | --- | --- |
| #1 | [mh Neoplasms] | 124145 |
| #2 | (adenocarcinoma* OR cancer* OR carcinoma* OR choricarcinoma* OR malignan* OR metasta* OR neoplasm* OR sarcoma* OR teratoma* OR tumo?r*):ti,ab,kw | 278528 |
| #3 | {OR #1-#2} | 289925 |
| #4 | [mh Malnutrition] | 6094 |
| #5 | [mh "Muscular Atrophy"] | 1424 |
| #6 | [mh "Weight Loss"] | 9011 |
| #7 | (cachex* OR malnourish* OR malnutri* OR (musc* NEXT (atroph* or loss)) OR sarcopeni* OR underweight OR wasting OR (weight NEAR/2 (loss OR redu*))):ti,ab,kw | 43678 |
| #8 | {OR #4-#7} | 47516 |
| #9 | [mh ^"Blood Glucose"] AND (fasting OR toleran*):ti,ab,kw | 9478 |
| #10 | [mh "Glucose Clamp Technique"] | 1161 |
| #11 | [mh "Insulin Resistance"] | 9615 |
| #12 | (((bloodglucose OR glucose) NEAR/2 (fasting OR toleran*)) OR (euglyc?emic NEAR/2 clamp*) OR homa-ir OR (insulin NEAR/2 (resistan* OR sensitiv*)) OR ogtt OR ((plasma* OR serum) NEXT insulin)):ti,ab,kw | 43105 |
| #13 | {OR #9-#12} | 47102 |
| #14 | #3 AND #8 AND #13 | 656 |

Revisiting insulin resistance in human cancer cachexia – a systematic review and meta-analysis

Journal of Cachexia, Sarcopenia and Muscle

Authors

Jonas Sørensena, Anna Hammershøia, Joan Miguel Màrmolb, Louise Lang Lehrskovc, d, Ole Nørgaarde, and Lykke Sylowa*.

Affiliations

a.	Department of Biomedical Sciences, Faculty of Health and Medical Sciences, University of Copenhagen, Copenhagen, Denmark

b.	Department of Nutrition, Exercise, and Sports, Faculty of Science, University of Copenhagen, Copenhagen, Denmark

c.	Center for Physical Activity Research (CFAS), Centre for Cancer and Organ Diseases, Copenhagen University Hospital – Rigshospitalet, Copenhagen, Denmark

d.	Department of Oncology, Copenhagen University Hospital – Herlev and Gentofte, Herlev, Denmark

e.	Department of Education, Danish Diabetes Knowledge Center, Copenhagen University Hospital – Steno Diabetes Center Copenhagen, Herlev, Denmark
