## Supplementary material for "Revisiting insulin resistance in human cancer cachexia – a systematic review and meta-analysis": S3

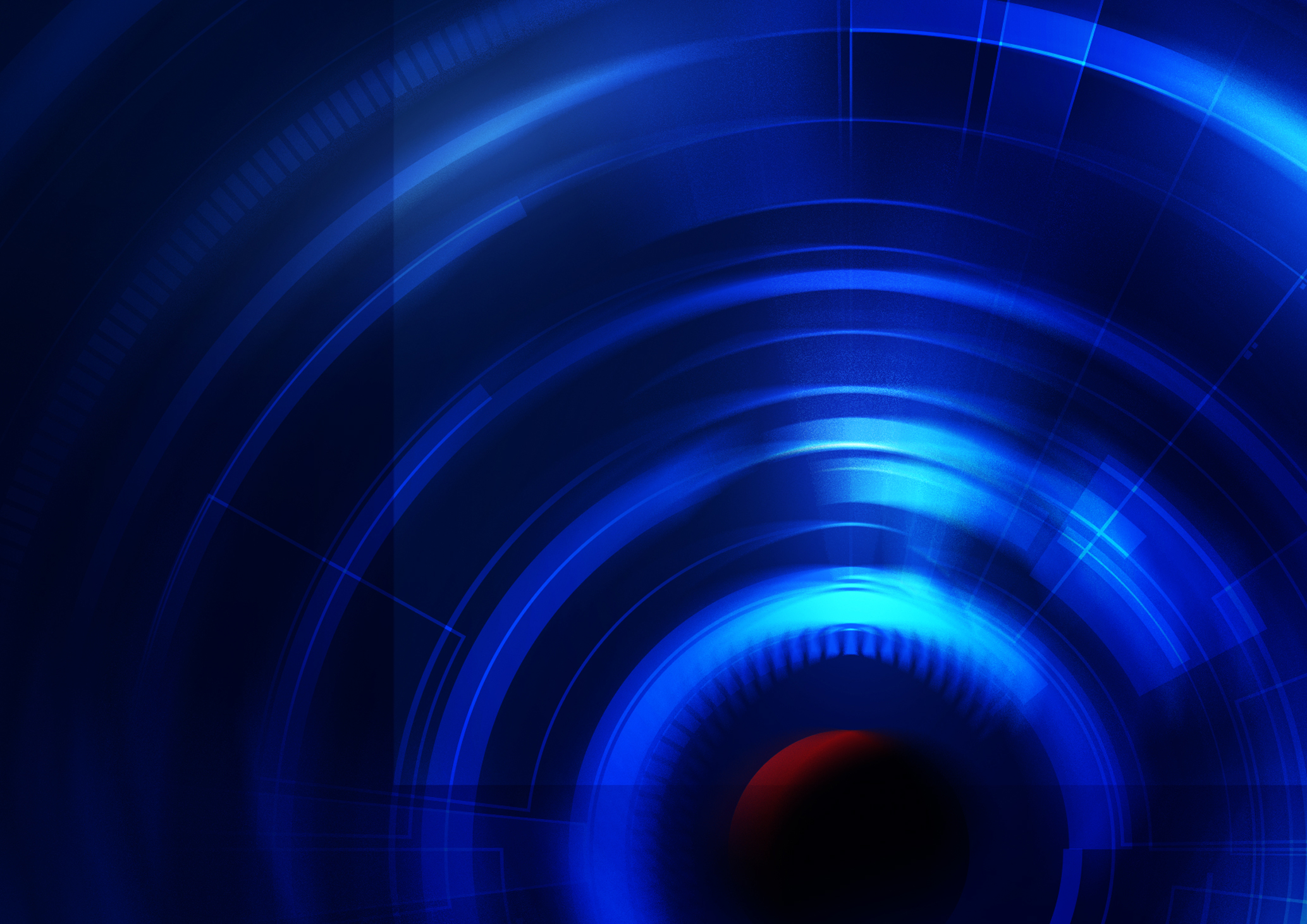

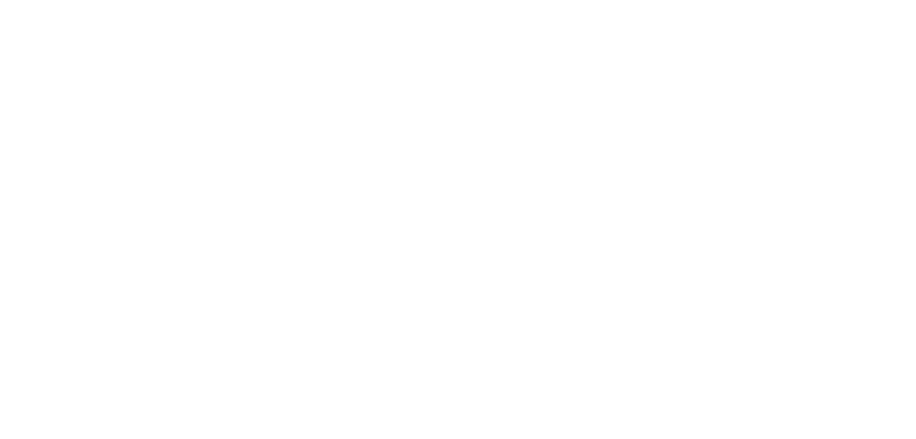

JBI CRITICAL APPRAISAL TOOL

**JBI checklist for
quasi-experimental
studies**

2023

introduction

**jbi.global**

CRICOS Provider Number 00123M

JBI is a global organization promoting and supporting evidence-based decisions that improve

health and health service delivery. JBI offers a unique range of solutions to access, appraise and apply the best available evidence, servicing over 90 countries. Working with 80+ universities, hospitals and NGOs from across the globe through the JBI Collaboration, JBI is a recognized global leader in evidence-based health care.

**JBI Systematic Reviews**

The core of evidence synthesis is the systematic review of literature of a particular intervention, condition or issue. The systematic review is essentially an analysis of the available evidence and a judgment of the effectiveness or otherwise of a practice, involving a series of complex steps. JBI takes a particular view on what counts as evidence and the methods utilized to synthesize those different types of evidence. In line with this broader view of evidence, JBI has developed theories, methodologies and rigorous processes for the critical appraisal and synthesis of these diverse forms of evidence in order to aid in clinical decision-making in health care. Guidance now exists for conducting reviews of effectiveness research, qualitative research, prevalence/incidence, etiology/risk, economic evaluations, text/opinion, diagnostic test accuracy, mixed-methods, umbrella reviews and scoping reviews. Further information regarding JBI systematic reviews can be found in the JBI Manual for Evidence Synthesis.

**JBI Critical Appraisal Tools**

All systematic reviews incorporate a process of critique or appraisal of the research evidence. The purpose of this appraisal for quantitative evidence is to determine the extent to which a study has addressed the possibility of bias in its design, conduct and analysis. All papers selected for inclusion in the systematic review (that is – those that meet the inclusion criteria described in the protocol) need to be subjected to rigorous appraisal by two critical appraisers. The results of this appraisal can then be used to inform synthesis and interpretation of the results of the study. Although designed for use in systematic reviews, JBI critical appraisal tools can also be used when creating Critically Appraised Topics, in journal clubs and as an educational tool.

**How were these tools developed?**

JBI critical appraisal tools have been developed by JBI and collaborators. The particular iteration of this tool was developed by the JBI Effectiveness Methodology Group following oversight by the JBI Scientific Committee.

Like the previous versions of these tools, this version presents signaling questions to prompt reviewers to identify whether certain safeguards of bias have been met, in the primary literature under review. However, unlike previous iterations of this tool, this version has separated questions into whether they provide an answer relating to internal, external or statistical conclusion validity. For questions related to internal validity, these have been further separated to identify what domain of bias they are referring. Finally, this tool has also been structured to facilitate judgments related to bias at different levels (e.g. bias at the outcome level or bias at the result level) where appropriate.

These tools have been approved following extensive peer review by the JBI Scientific Committee.

**How to cite**

Please use the following when citing this tool: Barker TH, Habibi N, Aromataris E, Stone JC, Leonardi-Bee J, Sears K, et al. The revised JBI critical appraisal tool for the assessment of risk of bias quasi-experimental studies. JBI Evid Synth. 2024;22(3):378-88.

| **RoB Assessor: A.H. and J.S.** | | **Date of Appraisal: 13.03.25** | | **Record Number: doi: 10.1042/CS19990273** | | | | |
| --- | --- | --- | --- | --- | --- | --- | --- | --- |
| **Study Author: Barber MD et al.** | | **Study Title: Metabolic response to feeding in weight-losing pancreatic cancer patients and its modulation by a ﬁsh-oil-enriched nutritional supplement** | | **Study Year: 2000** | | | | |
| **Internal Validity** | | | **Choice - Comments/Justification** | | **Yes** | **No** | **Unclear** | **N/A** |
| **Bias related to temporal precedence** | | | | | | | | |
| **1** | **Is it clear in the study what is the “cause” and what is the “effect” (i.e. there is no confusion about which variable comes first)?** | |  | | ☒ | ☐ | ☐ | ☐ |
| **Bias related to selection and allocation** | | | | | | | | |
| **2** | **Was there a control group?** | | Yes | | ☒ | ☐ | ☐ | ☐ |
| **Bias related to confounding factors** | | | | | | | | |
| **3** | **Were participants included in any comparisons similar?** | | Yes, 16 cancer patients with unequivocal diagnosis of pancreatic cancer who were losing weight. | | ☒ | ☐ | ☐ | ☐ |
| **Bias related to administration of intervention/exposure** | | | | | | | | |
| **4** | **Were the participants included in any comparisons receiving similar treatment/care, other than the exposure or intervention of interest?** | | The study is conducted before patients start cancer treatment | | ☐ | ☒ | ☐ | ☐ |

| **Bias related to assessment, detection and measurement of the outcome** | | | | | | |
| --- | --- | --- | --- | --- | --- | --- |
| **5** | **Were there multiple measurements of the outcome, both pre and post the intervention/exposure?** |  | **Yes** | **No** | **Unclear** | **N/A** |
|  | **Outcome 1** | Fasting levels of insulin and glucose at baseline measured by a blood test at day 1 of the study for all study participants | ☐ | ☒ | ☐ | ☐ |
|  | **Outcome 2** |  | ☐ | ☐ | ☐ | ☐ |
|  | **Outcome 3** |  | ☐ | ☐ | ☐ | ☐ |
|  | **Outcome 4** |  | ☐ | ☐ | ☐ | ☐ |
|  | **Outcome 5** |  | ☐ | ☐ | ☐ | ☐ |
|  | **Outcome 6** |  | ☐ | ☐ | ☐ | ☐ |
|  | **Outcome 7** |  | ☐ | ☐ | ☐ | ☐ |
| **6** | **Were the outcomes of participants included in any comparisons measured in the same way?** |  | **Yes** | **No** | **Unclear** | **N/A** |
|  | **Outcome 1** | Fasting levels of insulin and glucose at baseline measured by a blood test at day 1 of the study for all study participants | ☒ | ☐ | ☐ | ☐ |
|  | **Outcome 2** |  | ☐ | ☐ | ☐ | ☐ |
|  | **Outcome 3** |  | ☐ | ☐ | ☐ | ☐ |
|  | **Outcome 4** |  | ☐ | ☐ | ☐ | ☐ |
|  | **Outcome 5** |  | ☐ | ☐ | ☐ | ☐ |
|  | **Outcome 6** |  | ☐ | ☐ | ☐ | ☐ |
|  | **Outcome 7** |  | ☐ | ☐ | ☐ | ☐ |
| **7** | **Were outcomes measured in a reliable way?** |  | **Yes** | **No** | **Unclear** | **N/A** |
|  | **Outcome 1** | Fasting levels of insulin and glucose at baseline measured by a blood test at day 1 of the study for all study participants | ☒ | ☐ | ☐ | ☐ |
|  | **Outcome 2** |  | ☐ | ☐ | ☐ | ☐ |
|  | **Outcome 3** |  | ☐ | ☐ | ☐ | ☐ |
|  | **Outcome 4** |  | ☐ | ☐ | ☐ | ☐ |
|  | **Outcome 5** |  | ☐ | ☐ | ☐ | ☐ |
|  | **Outcome 6** |  | ☐ | ☐ | ☐ | ☐ |
|  | **Outcome 7** |  | ☐ | ☐ | ☐ | ☐ |

| **Bias related to participant retention** | | | | | | | | | | | | | | | |
| --- | --- | --- | --- | --- | --- | --- | --- | --- | --- | --- | --- | --- | --- | --- | --- |
| **8** | **Was follow-up complete and if not, were differences between groups in terms of their follow-up adequately described and analyzed?** | | | | | |  | |  | | | | | | |
|  | **Outcome 1** | | | | | | Follow up not relevant for outcome 1 | | **Yes** | **No** | | **Unclear** | | **N/A** | |
|  |  | Result 1 | | | | |  | | ☐ | ☐ | ☐ | | | ☐ | |
|  |  | Result 2 | | | | |  | | ☐ | ☐ | ☐ | | | ☐ | |
|  |  | Result 3 | | | | |  | | ☐ | ☐ | ☐ | | | ☐ | |
|  | **Outcome 2** | | | | | |  | | **Yes** | **No** | **Unclear** | | | **N/A** | |
|  |  | Result 1 | | | | |  | | ☐ | ☐ | ☐ | | | ☐ | |
|  |  | Result 2 | | | | |  | | ☐ | ☐ | ☐ | | | ☐ | |
|  |  | Result 3 | | | | |  | | ☐ | ☐ | ☐ | | | ☐ | |
|  | **Outcome 3** | | | | | |  | | **Yes** | **No** | **Unclear** | | | **N/A** | |
|  |  | Result 1 | | | | |  | | ☐ | ☐ | ☐ | | | ☐ | |
|  |  | Result 2 | | | | |  | | ☐ | ☐ | ☐ | | | ☐ | |
|  |  | Result 3 | | | | |  | | ☐ | ☐ | ☐ | | | ☐ | |
|  | **Outcome 4** | | | | | |  | | **Yes** | **No** | **Unclear** | | | **N/A** | |
|  |  | Result 1 | | | | |  | | ☐ | ☐ | ☐ | | | ☐ | |
|  |  | Result 2 | | | | |  | | ☐ | ☐ | ☐ | | | ☐ | |
|  |  | Result 3 | | | | |  | | ☐ | ☐ | ☐ | | | ☐ | |
|  | **Outcome 5** | | | | | |  | | **Yes** | **No** | **Unclear** | | | **N/A** | |
|  |  | Result 1 | | | | |  | | ☐ | ☐ | ☐ | | | ☐ | |
|  |  | Result 2 | | | | |  | | ☐ | ☐ | ☐ | | | ☐ | |
|  |  | Result 3 | | | | |  | | ☐ | ☐ | ☐ | | | ☐ | |
|  | **Outcome 6** | | | | | |  | | **Yes** | **No** | **Unclear** | | | **N/A** | |
|  |  | Result 1 | | | | |  | | ☐ | ☐ | ☐ | | | ☐ | |
|  |  | Result 2 | | | | |  | | ☐ | ☐ | ☐ | | | ☐ | |
|  |  | Result 3 | | | | |  | | ☐ | ☐ | ☐ | | | ☐ | |
|  | **Outcome 7** | | | | | |  | | **Yes** | **No** | **Unclear** | | | **N/A** | |
|  |  | Result 1 | | | | |  | | ☐ | ☐ | ☐ | | | ☐ | |
|  |  | Result 2 | | | | |  | | ☐ | ☐ | ☐ | | | ☐ | |
|  |  | Result 3 | | | | |  | | ☐ | ☐ | ☐ | | | ☐ | |
|  | **Statistical Conclusion Validity** | | | | | | | |  |  |  | | |  | |
| **9** | **Was appropriate statistical analysis used?** | | | | | | |  |  | | | | | | |
|  | **Outcome 1** | | |  | | | | Statical analysis not relevant for outcome 1 | **Yes** | **No** | **Unclear** | | | **N/A** | |
|  |  | Result 1 | | | | | |  | ☐ | ☐ | ☐ | | | ☐ | |
|  |  | Result 2 | | | | | |  | ☐ | ☐ | ☐ | | | ☐ | |
|  |  | Result 3 | | | | | |  | ☐ | ☐ | ☐ | | | ☐ | |
|  | **Outcome 2** | | |  | | | |  | **Yes** | **No** | **Unclear** | | | **N/A** | |
|  |  | Result 1 | | | | | |  | ☐ | ☐ | ☐ | | | ☐ | |
|  |  | Result 2 | | | | | |  | ☐ | ☐ | ☐ | | | ☐ | |
|  |  | Result 3 | | | | | |  | ☐ | ☐ | ☐ | | | ☐ | |
|  | **Outcome 3** | | |  | | | |  | **Yes** | **No** | **Unclear** | | | **N/A** | |
|  |  | Result 1 | | | | | |  | ☐ | ☐ | ☐ | | | ☐ | |
|  |  | Result 2 | | | | | |  | ☐ | ☐ | ☐ | | | ☐ | |
|  |  | Result 3 | | | | | |  | ☐ | ☐ | | | ☐ | | ☐ |
|  | **Outcome 4** | | | | | | |  | **Yes** | **No** | | | **Unclear** | | **N/A** |
|  |  | Result 1 | | | | | |  | ☐ | ☐ | | | ☐ | | ☐ |
|  |  | Result 2 | | | | | |  | ☐ | ☐ | | | ☐ | | ☐ |
|  |  | Result 3 | | | | | |  | ☐ | ☐ | | | ☐ | | ☐ |
|  | **Outcome 5** | | | | | | |  | **Yes** | **No** | | | **Unclear** | | **N/A** |
|  |  | Result 1 | | | | | |  | ☐ | ☐ | | | ☐ | | ☐ |
|  |  | Result 2 | | | | | |  | ☐ | ☐ | | | ☐ | | ☐ |
|  |  | Result 3 | | | | | |  | ☐ | ☐ | | | ☐ | | ☐ |
|  | **Outcome 6** | | | | | | |  | **Yes** | **No** | | | **Unclear** | | **N/A** |
|  |  | Result 1 | | | | | |  | ☐ | ☐ | | | ☐ | | ☐ |
|  |  | Result 2 | | | | | |  | ☐ | ☐ | | | ☐ | | ☐ |
|  |  | Result 3 | | | | | |  | ☐ | ☐ | | | ☐ | | ☐ |
|  | **Outcome 7** | | | | | | |  | **Yes** | **No** | | | **Unclear** | | **N/A** |
|  |  | Result 1 | | | | | |  | ☐ | ☐ | | | ☐ | | ☐ |
|  |  | Result 2 | | | | | |  | ☐ | ☐ | | | ☐ | | ☐ |
|  |  | Result 3 | | | | | |  | ☐ | ☐ | | | ☐ | | ☐ |
| **Overall appraisal:** | | | **Include: ☒** | | **Exclude: ☐** | **Seek Further Info: ☐** | | | | | | | | | |
| **Comments:**  **For our review, we only extract data about fasting insulin and glucose levels at baseline for patients, and this study is appraised according to measurement of these parameters.** | | | | | | | | | | | | | | | |

© JBI, 2022. All rights reserved. JBI grants use of these

 tools for research purposes only. All other enquiries

 should be sent to 
