## Supplementary material for "Revisiting insulin resistance in human cancer cachexia – a systematic review and meta-analysis": S10

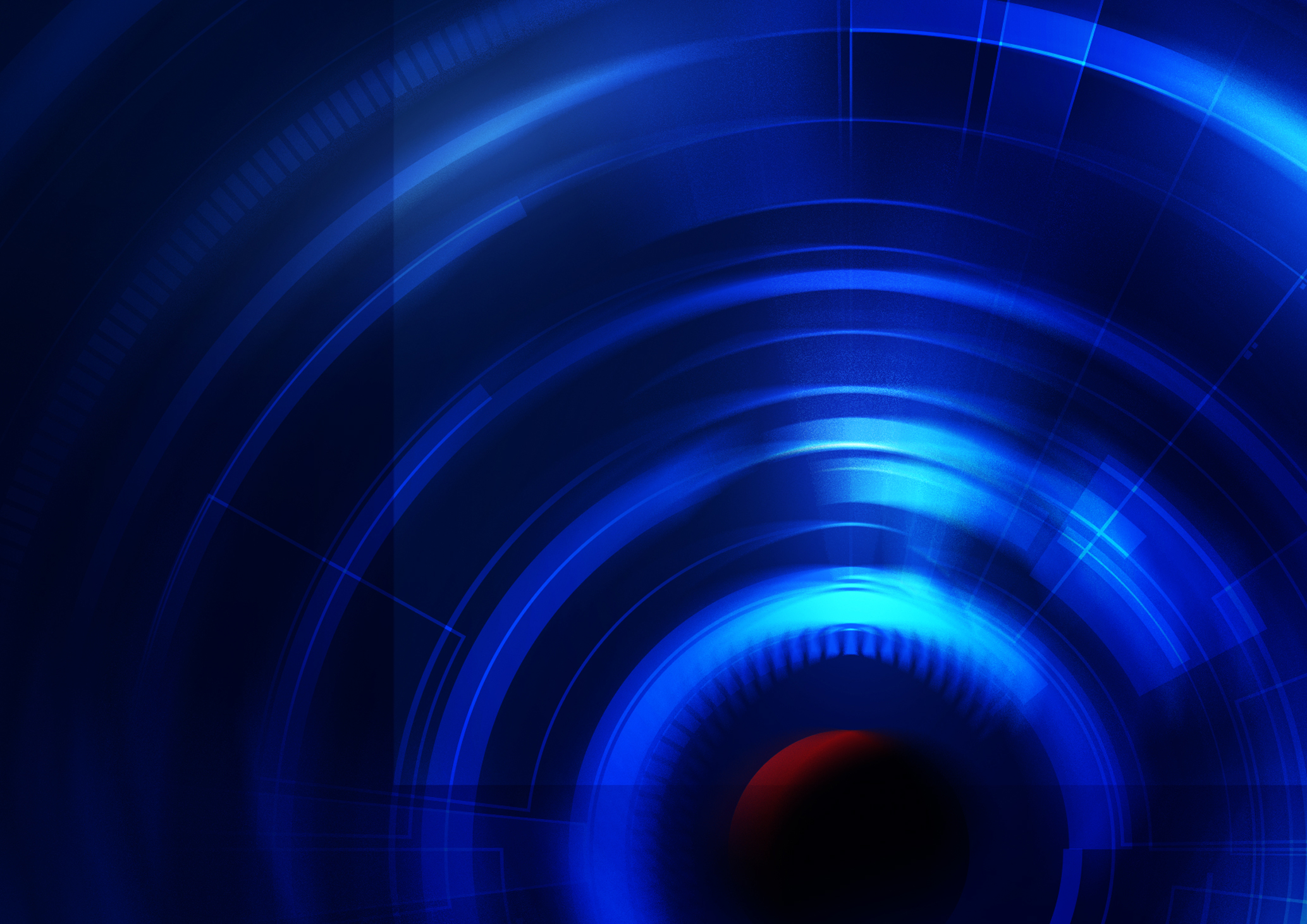

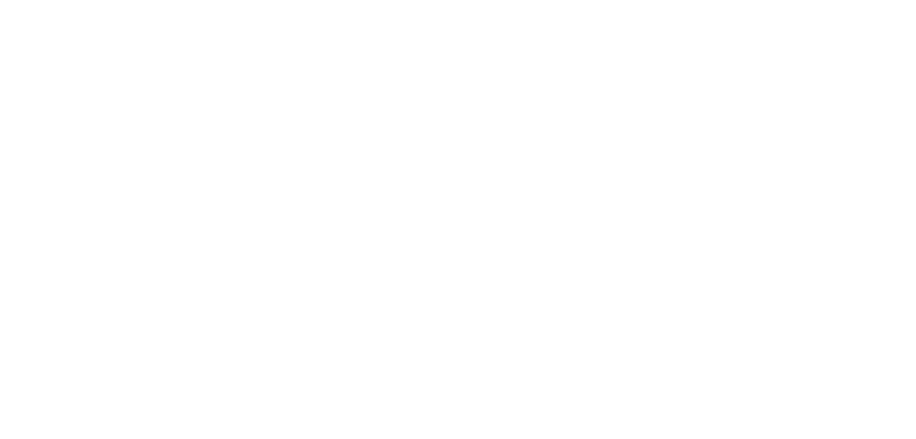

JBI CRITICAL APPRAISAL TOOL

**JBI checklist for
quasi-experimental
studies**

2023

introduction

**jbi.global**

CRICOS Provider Number 00123M

JBI is a global organization promoting and supporting evidence-based decisions that improve

health and health service delivery. JBI offers a unique range of solutions to access, appraise and apply the best available evidence, servicing over 90 countries.

| **RoB Assessor: A.H. and J.S.** | | **Date of Appraisal: 14.03.25** | | **Record Number: doi.org/10.1002/1097-0142(19850101)55:1+<225::aid-cncr2820551304>3.0.co;2-7** | | | | |
| --- | --- | --- | --- | --- | --- | --- | --- | --- |
| **Study Author: Herber D et al.** | | **Study Title: Metabolic abnormalities in the cancer patient** | | **Study Year: 1985** | | | | |
| **Internal Validity** | | | **Choice - Comments/Justification** | | **Yes** | **No** | **Unclear** | **N/A** |
| **Bias related to temporal precedence** | | | | | | | | |
| **1** | **Is it clear in the study what is the “cause” and what is the “effect” (i.e. there is no confusion about which variable comes first)?** | |  | | ☒ | ☐ | ☐ | ☐ |
| **Bias related to selection and allocation** | | | | | | | | |
| **2** | **Was there a control group?** | |  | | ☒ | ☐ | ☐ | ☐ |
| **Bias related to confounding factors** | | | | | | | | |
| **3** | **Were participants included in any comparisons similar?** | | Yes, weight-losing cancer patients | | ☒ | ☐ | ☐ | ☐ |
| **Bias related to administration of intervention/exposure** | | | | | | | | |
| **4** | **Were the participants included in any comparisons receiving similar treatment/care, other than the exposure or intervention of interest?** | | It is unclear if the patients had received any cancer treatment before the study. | | ☐ | ☐ | ☒ | ☐ |
