## Supplementary material for "Revisiting insulin resistance in human cancer cachexia – a systematic review and meta-analysis": S14

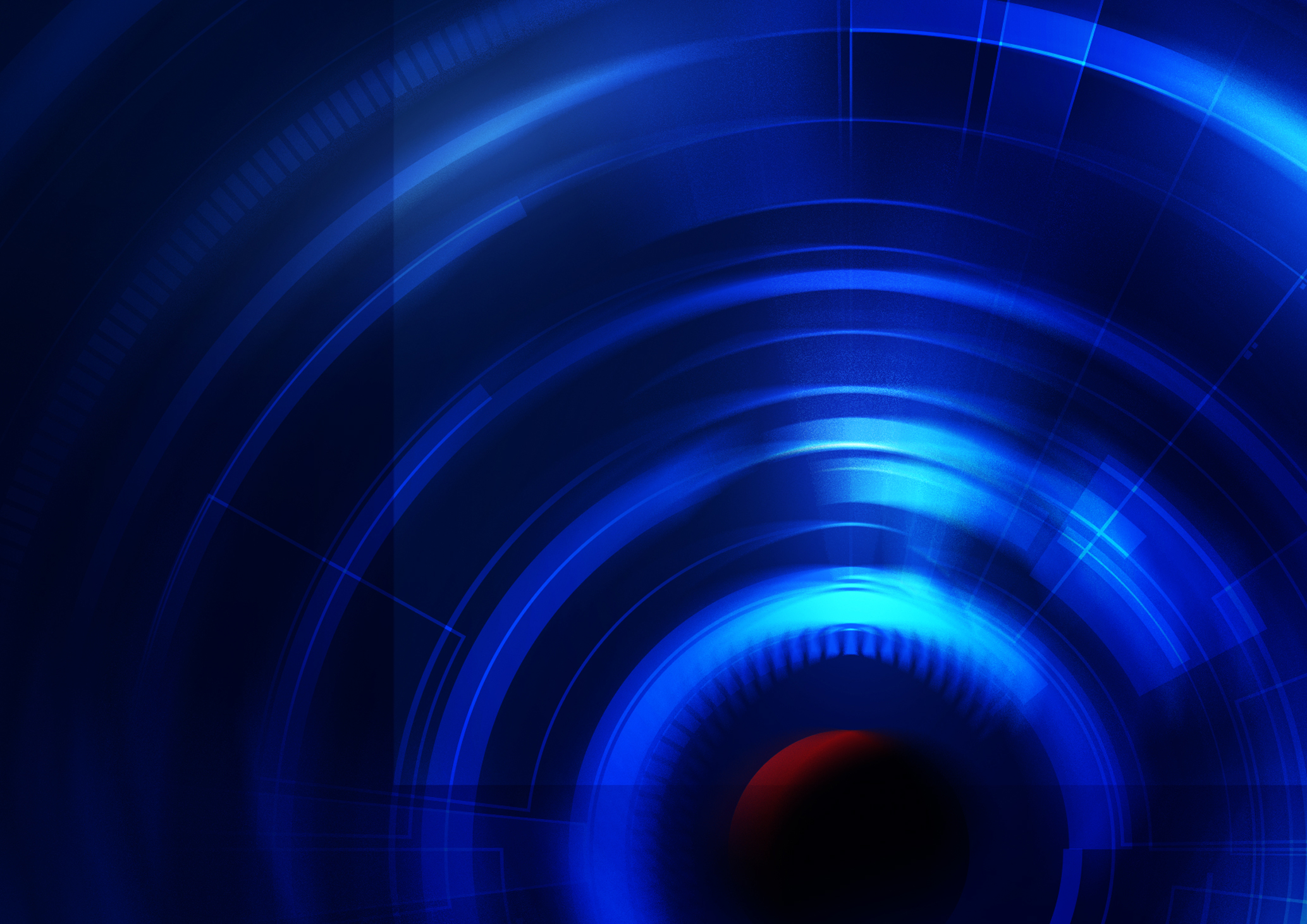

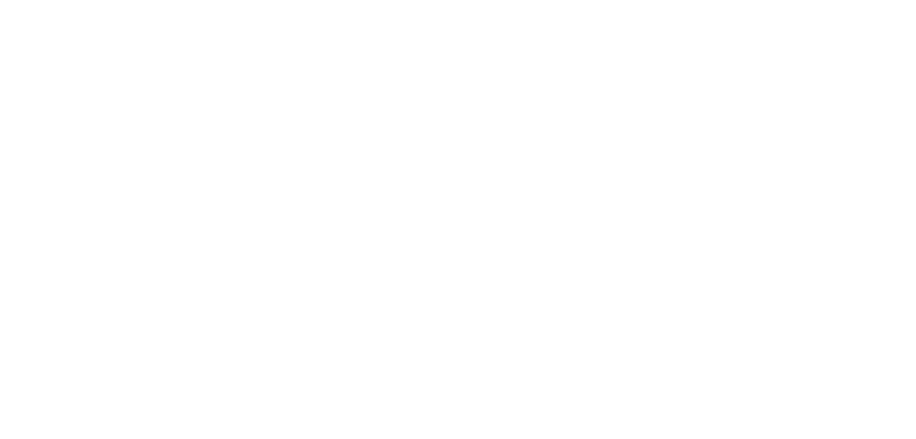

JBI CRITICAL APPRAISAL TOOL

**JBI checklist for
quasi-experimental
studies**

2023

introduction

**jbi.global**

CRICOS Provider Number 00123M

JBI is a global organization promoting and supporting evidence-based decisions that improve

health and health service delivery. JBI offers a unique range of solutions to access, appraise and apply the best available evidence, servicing over 90 countries.
