## Supplementary material for "Revisiting insulin resistance in human cancer cachexia – a systematic review and meta-analysis": S20

**Table S20: Excluded reports after full-text screening**

| **Report** | **Reason for exclusion** |
| --- | --- |
| Allen S, Brown V, White D, King D, Hunt J, Prabhu P, Rockall T, Preston S, Sultan J. Multi-modal prehabilitation during neoadjuvant therapy prior to resection for oesophagogastric cancer: a pilot randomised controlled trial. Br J Surg. 2019;106:90‐1. | No fasting glucose/insulin data |
| Barber MD, Fearon KC, Tisdale MJ, McMillan DC, Ross JA. Effect of a fish oil-enriched nutritional supplement on metabolic mediators in patients with pancreatic cancer cachexia. Nutr Cancer. 2001;40(2):118-24. | No fasting glucose/insulin data |
| Barber MD, McMillan DC, Wallace AM, Ross JA, Preston T, Fearon KC. The response of leptin, interleukin-6 and fat oxidation to feeding in weight-losing patients with pancreatic cancer. Br J Cancer. 2004 Mar 22;90(6):1129-32. | Other reason (uses subgroup from other study) |
| Begenik H, Aslan M, Dulger AC, Emre H, Kemik A, Kemik O, Esen R. Serum leptin levels in gastric cancer patients and the relationship with insulin resistance. Arch Med Sci. 2015 Apr 25;11(2):346-52. | No weight loss data |
| Bennegård K, Lindmark L, Edén E, Svaninger G, Lundholm K. Flux of amino acids across the leg in weight-losing cancer patients. Cancer Res. 1984 Jan;44(1):386-93. | Other reason (uses subgroup from other study) |
| Byerley LO, Heber D, Bergman RN, Dubria M, Chi J. Insulin action and metabolism in patients with head and neck cancer. Cancer. 1991 Jun 1;67(11):2900-6. | No weight loss data |
| Caeiro L, Anderson LJ, Liu H, Krumm K, Garcia JM. THU512 Elevated insulin like growth factor binding protein 2 levels are associated with muscle wasting and increased insulin sensitivity in cancer patients. J Endocr Soc. 2023 Nov 1;7(Suppl 1):bvad114.2140. | No weight loss data |
| Chlebowski RT, Heber D, Richardson B, Block JB. Influence of hydrazine sulfate on abnormal carbohydrate metabolism in cancer patients with weight loss. Cancer Res. 1984 Feb;44(2):857-61. | No fasting glucose/insulin data |
| Copeland GP, Al-Sumidaie AM, Leinster SJ, Davis JC, Hipkin LH. Glucose metabolism in patients with gastrointestinal malignancy but without excessive weight loss. Eur J Surg Oncol. 1987 Feb;13(1):11-6. | No fasting glucose/insulin data |
| Copeland GP, Leinster SJ, Davis JC, Hipkin LH. Postoperative glucose metabolism in patients with gastrointestinal malignancy. Eur J Surg Oncol. 1988 Dec;14(6):677-83. | Not cancer |
| Dawson JK, Dorff TB, Todd Schroeder E, Lane CJ, Gross ME, Dieli-Conwright CM. Impact of resistance training on body composition and metabolic syndrome variables during androgen deprivation therapy for prostate cancer: a pilot randomized controlled trial. BMC Cancer. 2018 Apr 3;18(1):368. | No weight loss data |
| de Carvalho TM, Miguel Marin D, da Silva CA, de Souza AL, Talamoni M, Lima CS, Monte Alegre S. Evaluation of patients with head and neck cancer performing standard treatment in relation to body composition, resting metabolic rate, and inflammatory cytokines. Head Neck. 2015 Jan;37(1):97-102. | No weight loss data |
| Dong J, Zeng Y, Zhang P, Li C, Chen Y, Li Y, Wang K. Serum IGFBP2 Level Is a New Candidate Biomarker of Severe Malnutrition in Advanced Lung Cancer. Nutr Cancer. 2020;72(5):858-63. | No weight loss data |
| Dülger H, Alici S, Sekeroğlu MR, Erkog R, Ozbek H, Noyan T, Yavuz M. Serum levels of leptin and proinflammatory cytokines in patients with gastrointestinal cancer. Int J Clin Pract. 2004 Jun;58(6):545-9. | No weight loss data |
| Fouladiun M, Körner U, Bosaeus I, Daneryd P, Hyltander A, Lundholm KG. Body composition and time course changes in regional distribution of fat and lean tissue in unselected cancer patients on palliative care--correlations with food intake, metabolism, exercise capacity, and hormones. Cancer. 2005 May 15;103(10):2189-98. | No fasting glucose/insulin data |
| Gambardella A, Paolisso G, D'Amore A, Granato M, Verza M, Varricchio M. Different contribution of substrates oxidation to insulin resistance in malnourished elderly patients with cancer. Cancer. 1993 Nov 15;72(10):3106-13. | No fasting glucose/insulin data |
| Garcia JM, Garcia-Touza M, Hijazi RA, Taffet G, Epner D, Mann D, Smith RG, Cunningham GR, Marcelli M. Active ghrelin levels and active to total ghrelin ratio in cancer-induced cachexia. J Clin Endocrinol Metab. 2005 May;90(5):2920-6. | Other reason (includes patients with diabetes) |
| Iwai N, Sakai H, Oka K, Sakagami J, Okuda T, Hattori C, Taniguchi M, Hara T, Tsuji T, Komaki T, Kagawa K, Doi T, Ishikawa T, Yasuda H, Itoh Y. Predictors of response to anamorelin in gastrointestinal cancer patients with cachexia: a retrospective study. Support Care Cancer. 2023 Jan 14;31(2):115. | No weight loss data |
| Kerem M, Ferahkose Z, Yilmaz UT, Pasaoglu H, Ofluoglu E, Bedirli A, Salman B, Sahin TT, Akin M. Adipokines and ghrelin in gastric cancer cachexia. World J Gastroenterol. 2008 Jun 21;14(23):3633-41. | Other reason (includes patients with diabetes) |
| Kokal WA, McCulloch A, Wright PD, Johnston ID. Glucose turnover and recycling in colorectal carcinoma. Ann Surg. 1983 Nov;198(5):601-4. | No fasting glucose/insulin data |
| Körber J, Pricelius S, Heidrich M, Müller MJ. Increased lipid utilization in weight losing and weight stable cancer patients with normal body weight. Eur J Clin Nutr. 1999 Sep;53(9):740-5. | No fasting glucose/insulin data |
| Li R, Ma ML, Song YY, Pei J, Zhong RB, Qian JL, Yan B, Zhang XY, Shen J, Han BH. Investigation on the nutrition status of 132 advanced lung cancer patients in primary treatment. Tumor. 2008;28(4):353-6. | No weight loss data |
| Liao WC, Chen PR, Huang CC, Chang YT, Huang BS, Chang CC, Wu MS, Chow LP. Relationship between pancreatic cancer-associated diabetes and cachexia. J Cachexia Sarcopenia Muscle. 2020 Aug;11(4):899-908. | No fasting glucose/insulin data |
| Lyu H, Yang X, Ding R, Cui H, Qiao J, Zhu M, Wei J. A prospective observational study on nutritional status of patients with pancreatic tumor. Chin J Clin Nutr. 2017;25(2):94-8. | No weight loss data |
| Makino T, Noguchi Y, Yoshikawa T, Doi C, Nomura K. Circulating interleukin 6 concentrations and insulin resistance in patients with cancer. Br J Surg. 1998 Dec;85(12):1658-62. | No fasting glucose/insulin data |
| Mendes MCS, Juliani FL, Branbilla SR, Souza AL, Alegre SM, Costa FO, Martinez CAR, Carvalheira JBC. Body composition and insulin sensitivity in patients with rectal cancer. Hematol Transfus Cell Ther. 2024;46(Suppl 2):S5-6. | No weight loss data |
| Newman E, Heslin MJ, Wolf RF, Pisters PW, Brennan MF. The effect of insulin on glucose and protein metabolism in the forearm of cancer patients. Surg Oncol. 1992 Aug;1(4):257-67. | No fasting glucose/insulin data |
| Rantaniemi L, Siltari A, Harju E, Murtola TJ. Can supervised exercise impact on metabolic markers and physical activity during androgen-deprivation therapy in prostate cancer patients? - randomized controlled pilot trial. Cancer Res. 2022;82(Suppl 12):CT171. | No weight loss data |
| Ruan GT, Deng L, Xie HL, Shi JY, Liu XY, Zheng X, Chen Y, Lin SQ, Zhang HY, Liu CA, Ge YZ, Song MM, Hu CL, Zhang XW, Yang M, Hu W, Cong MH, Zhu LC, Wang KH, Shi HP. Systemic inflammation and insulin resistance-related indicator predicts poor outcome in patients with cancer cachexia. Cancer Metab. 2024 Jan 25;12(1):3. | No fasting glucose/insulin data |
| Shaw JH, Humberstone DA, Holdaway C. Weight loss in patients with head and neck cancer: malnutrition or tumour effect? Aust N Z J Surg. 1988 Jun;58(6):505-9. | No weight loss data |
| Smiechowska J, Utech A, Taffet G, Hayes T, Marcelli M, Garcia JM. Adipokines in patients with cancer anorexia and cachexia. J Investig Med. 2010 Mar;58(3):554-9. | Other reason (uses subgroup from other study) |
| Tayek JA, Bulcavage L, Chlebowski RT. Relationship of hepatic glucose production to growth hormone and severity of malnutrition in a population with colorectal carcinoma. Cancer Res. 1990 Apr 1;50(7):2119-22. | No weight loss data |
| Tayek JA, Chlebowski RT. Metabolic response to chemotherapy in colon cancer patients. JPEN J Parenter Enteral Nutr. 1992 Nov-Dec;16(6 Suppl):65S-71S. | No weight loss data |
| Tayek JA, Manglik S, Abemayor E. Insulin secretion, glucose production, and insulin sensitivity in underweight and normal-weight volunteers, and in underweight and normal-weight cancer patients: a Clinical Research Center study. Metabolism. 1997 Feb;46(2):140-5. | No weight loss data |
| Tůma P, Hložek T, Kamišová J, Gojda J. Monitoring of circulating amino acids in patients with pancreatic cancer and cancer cachexia using capillary electrophoresis and contactless conductivity detection. Electrophoresis. 2021 Oct;42(19):1885-1891. | No weight loss data |
| Winter A, MacAdams J, Chevalier S. Normal protein anabolic response to hyperaminoacidemia in insulin-resistant patients with lung cancer cachexia. Clin Nutr. 2012 Oct;31(5):765-73. | No weight loss data |
| Yoshikawa T, Noguchi Y, Doi C, Makino T, Nomura K. Insulin resistance in patients with cancer: relationships with tumor site, tumor stage, body-weight loss, acute-phase response, and energy expenditure. Nutrition. 2001 Jul-Aug;17(7-8):590-3. | No weight loss data |
| Zwickl H, Hackner K, Köfeler H, Krzizek EC, Muqaku B, Pils D, Scharnagl H, Solheim TS, Zwickl-Traxler E, Pecherstorfer M. Reduced LDL-Cholesterol and Reduced Total Cholesterol as Potential Indicators of Early Cancer in Male Treatment-Naïve Cancer Patients With Pre-cachexia and Cachexia. Front Oncol. 2020 Aug 4;10:1262. | Wrong population |

Revisiting insulin resistance in human cancer cachexia – a systematic review and meta-analysis

Journal of Cachexia, Sarcopenia and Muscle

Authors

Jonas Sørensena, Anna Hammershøia, Joan Miguel Màrmolb, Louise Lang Lehrskovc, d, Ole Nørgaarde, and Lykke Sylowa*.

Affiliations

a.	Department of Biomedical Sciences, Faculty of Health and Medical Sciences, University of Copenhagen, Copenhagen, Denmark

b.	Department of Nutrition, Exercise, and Sports, Faculty of Science, University of Copenhagen, Copenhagen, Denmark

c.	Center for Physical Activity Research (CFAS), Centre for Cancer and Organ Diseases, Copenhagen University Hospital – Rigshospitalet, Copenhagen, Denmark

d.	Department of Oncology, Copenhagen University Hospital – Herlev and Gentofte, Herlev, Denmark

e.	Department of Education, Danish Diabetes Knowledge Center, Copenhagen University Hospital – Steno Diabetes Center Copenhagen, Herlev, Denmark
